# Multi-layered genetic approaches to identify approved drug targets

**DOI:** 10.1101/2023.03.21.23285637

**Authors:** Marie C. Sadler, Chiara Auwerx, Patrick Deelen, Zoltán Kutalik

## Abstract

Drugs targeting genes that harbor natural variations associated with the disease the drug is indicated for have increased odds to be approved. Various approaches have been proposed to identify likely causal genes for complex diseases, including gene-based genome-wide association studies (GWAS), rare variant burden tests in whole exome sequencing studies (Exome) or integration of GWAS with expression/protein quantitative trait loci (eQTL-GWAS/pQTL-GWAS). Here, we compare gene-prioritization approaches on 30 common clinical traits and benchmarked their ability to recover drug target genes defined using a combination of five drug databases. Across all traits, the top prioritized genes were enriched for drug targets with odds ratios (ORs) of 2.17, 2.04, 1.81 and 1.31 for the GWAS, eQTL-GWAS, Exome and pQTL-GWAS methods, respectively. We quantified the performance of these methods using the area under the receiver operating characteristic curve as metric, and adjusted for differences in testable genes and data origins. GWAS performed significantly better (54.3%) than eQTL (52.8%) and pQTL-GWAS (51.3%), but not significantly so against the Exome approach (51.7% *vs* 52.8% for GWAS restricted to UK Biobank data). Furthermore, our analysis showed increased performance when diffusing gene scores on gene networks. However, substantial improvements in the protein-protein interaction network may be due to circularity in the data generation process, leading to the node (gene) degree being the best predictor for drug target genes (OR = 8.7, 95% CI = 7.3-10.4) and warranting caution when applying this strategy. In conclusion, we systematically assessed strategies to prioritize drug target genes highlighting promises and potential pitfalls of current approaches.

## Introduction

Drugs whose targets have genetic support were found to be more likely to succeed in clinical trials [1, 2]. Although multiple methods have been proposed to establish such genetic support, leveraging genetic data to find disease genes and ultimately drug targets has proven to be challenging [3, 4, 5, 6]. The most straightforward approach maps genome-wide association studies (GWASs) signals to the closest genes with more sophisticated methods incorporating linkage disequilibrium (LD) structure and gene annotation information to compute gene scores [7, 8, 9]. Over the past decade, large-scale molecular quantitative trait loci (mQTL) datasets facilitated the discovery of disease mechanisms and the identification of potential new drug targets [10, 11, 12, 13, 14, 15]. Several methods, including Mendelian randomization studies, transcriptome-wide association studies and colocalization methods have integrated expression and protein QTL data with GWAS studies to pinpoint likely causal genes for complex traits and diseases [16, 17, 18, 19, 20, 21, 22]. More recently, the availability of high-throughput sequencing data enabled the discovery and analysis of rare variants and their aggregated effects to reveal gene-disease associations [23, 24]. Whole exome sequencing (WES) in the UK Biobank (UKBB) showed that genes prioritized this way are 3.6 times more likely to be targets of drugs approved by the Food and Drug Administration (FDA) [25].

Genes prioritised by GWASs, mQTL-GWAS integration methods and WES burden tests may not be drug targets themselves, but up- or downstream of those in pharmacological pathways. Propagating gene prioritization scores on networks has proven to be a promising approach to identify known drug target genes [26, 27, 28, 29, 30]. Starting from seed genes (i.e., prioritized disease-associated genes), network connectivity can identify neighbouring genes that strongly interact with disease genes, but lack a direct genetic evidence that explain their therapeutic effect. Gene networks can be derived from literature or high-throughput experiments and thus are prone to yield very different results when used for (seed) gene score diffusion [31].

Here, we took a comprehensive approach to examine the contribution of each method component to the success of drug target prioritization. First, we focused on four different approaches to **prioritise (seed) genes**: 1) LD-aware gene score computation from the largest GWASs with full publicly available summary statistics (Pascal [9]); 2) Mendelian Randomisation (MR) combining tissue-wide expression QTLs and GWAS (eQTL-GWAS); 3) MR combining plasma protein QTL with GWAS (pQTL-GWAS); 4) UKBB WES burden tests (Exome). We then used three different networks to **diffuse the seed gene scores**: 1) STRING protein-protein interaction (PPI) network [32]; 2) an RNA-seq co-expression network [33]; 3) the FAVA network [34]. All 12 combinations of the four seed generating methods and the three networks were applied to thirty traits (Figure 1) using five different reference sets of target genes (DrugBank [35], Ruiz *et al.* [36], ChEMBL [37], DGIdb [38] and STITCH [39]). Overall, we provide an in-depth comparison of all combinations of these approaches, identifying their respective strengths and caveats.

**Figure 1:**
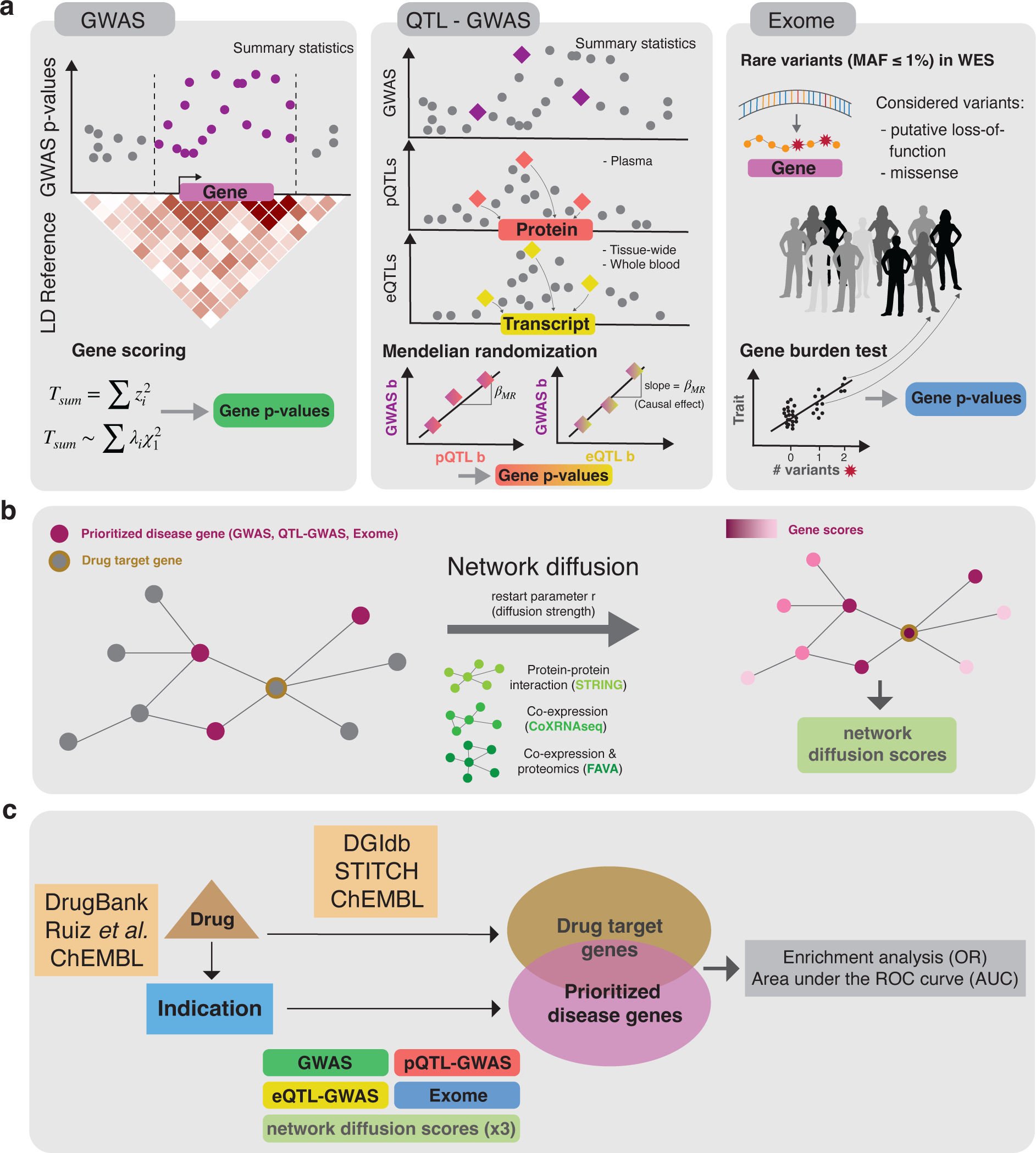
Overview of the analysis workflow. **a** Three different gene prioritization methods were tested in this study. The first one uses GWAS summary statistics as input (GWAS); the second combines molecular QTL and GWAS summary statistics (QTL-GWAS): either expression QTL (eQTL) or protein QTL (pQTL) data; the third leverages individual-level whole exome sequencing (WES) data (Exome). In the GWAS method, gene p-values are based on the sum of squared SNP z-scores (*T*_sum_) that fol-lows a weighted 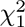 distribution. The QTL-GWAS method integrates QTL and GWAS summary statistics through Mendelian randomization (MR). MR causal effect sizes (*β*_MR_) are calculated from GWAS and mQTL effect sizes (GWAS b and mQTL b, respectively) and gene scores are the corresponding p-values. The Exome method aggregates rare variants from WES data. Putative loss-of-function and missense variants with minor allele frequencies (MAF) below 1% are collapsed in burden tests which results in gene p-values. The different approaches were benchmarked for their ability to prioritize drug target genes. **b** The effect of network diffusion using three different network types and different diffusion strengths (i.e. restart parameter *r*) was evaluated. Drug target genes may only be prioritized following signal propagation from neighboring disease genes. **c** Diseases were linked to target genes through public drug databases: first, we used drug-indication information to connect the 30 traits to drugs and then leveraged drug-target information to link the drugs to genes. Prioritized disease genes and corresponding diffusion scores (obtained via strategies described in panel **a** and **b**) were then tested for overlap with drug target genes through Fisher’s exact tests resulting in odds ratios (ORs) and through area under the receiver operating characteristic curve (AUC) values.

## Results

### Overview of the analysis

In this study, we calculated gene prioritization scores and tested their ability to identify drug targets across thirty traits (Figure 1). We focused on three types of methods termed *GWAS*, *QTL-GWAS* and *Exome* that allow the computation of gene scores provided genetic association data (Figure 1a).

The GWAS method takes as input GWAS summary statistics together with a matching LD reference panel. Gene p-values are calculated based on the sum of squared test statistics for SNPs falling into the gene region [9]. The QTL-GWAS methods integrate GWAS summary statistics with molecular QTL data for the gene of interest. We calculated gene scores using i) expression QTL data (*eQTL-GWAS*) from the largest available whole blood eQTL study (eQTLGen study, *n* = 31,684) as well as tissue-wide eQTL data from the GTEx consortium v8 (*n* = 65-573 for 48 tissue types) and ii) protein QTL data (*pQTL-GWAS*) from the largest available plasma pQTL study (deCODE study, *n* = 35,559) [14] and i. Integration was done by performing Mendelian randomization (MR) analyses using either the protein or transcript as exposure and the GWAS trait as outcome. If not specified otherwise, the eQTL-GWAS method refers to the tissue-wide analysis in which the eQTLGen and GTEx data are combined by considering the tissue for which the MR effect was the most significant (Methods). While the GWAS and QTL-GWAS methods focuse on common genetic variants, the Exome method considers only rare variants from WES data with minor allele frequencies (MAF) below 1%. Gene scores were based on gene burden tests that aggregate putative loss-of-function and missense variants, and we used the resulting p-values from the WES analysis in the UKBB [25]. To allow for a fair comparison with the Exome method while also exploiting disease-specific consortia GWAS summary statistics with maximized case counts, we calculated gene prioritization scores for the GWAS and QTL-GWAS methods using both consortia GWAS and UKBB GWAS data (Table S1-3; Methods).

Disease genes may not coincide with drug target genes, but they may be in close proximity in terms of molecular interaction (Figure 1b). Through diffusion based on random walks, we leveraged network connectivity to prioritize neighbours of disease genes, which may be drug targets. We tested this hypothesis on three different network types: the STRING PPI network which relies on literature interactions among other data types [32], a gene co-expression network based on 31,499 RNA-seq samples (*CoXRNAseq*) [33], and gene co-expression network based on single cell RNA-seq and proteomics data (*FAVA*) [34]. Gene prioritization scores were obtained following diffusion at six different restart parameter values (r = 0, 0.2, 0.4, 0.6, 0.8, 1) (Methods).

Disease drug target genes were defined using public databases. Specifically, drug-disease indication were retrieved from DrugBank [35], Ruiz *et al.* [36], and ChEMBL [37], while drug-drug target pairs originate from DGIdb [38], STITCH [39], and ChEMBL [37]. Drug target enrichment analyses were calculated for the following five database combinations: DrugBank/DGIdb, DrugBank/STITCH, Ruiz/DGIdb, Ruiz/STITCH, and ChEMBL/ChEMBL.

Finally, prioritized disease genes, defined as the top 1% of genes identified through the 12 combinations of gene prioritization and network diffusion methods (5% for combinations involving the pQTL-GWAS method to account for the smaller set of testable genes), were then tested for enrichment with the five drug target genes using Fisher’s exact test (Figure 1c). Background genes were defined as all genes that could be tested by the respective method, and sensitivity analyses were performed on background genes testable for all methods. Second, we calculated the area under the receiver operating characteristic curve (AUC) values which has the advantage of not requiring any thresholds. To compute a combined enrichment score per method, we aggregate results across traits and drug databases termed overall ORs or overall AUC values (Methods).

### Concordance of prioritized genes among gene scoring methods

We first analyzed whether genes prioritized by the GWAS, QTL-GWAS and Exome methods were concordant (Figure 2). For each of the thirty traits (Figure 2a), we calculated gene scores for the testable autosomal protein-coding genes (GWAS: *∼*19,150, eQTL-GWAS: *∼*12,550 (blood) and *∼*16,250 (tissue-wide), pQTL-GWAS: *∼*1,870, Exome: *∼*18,800). In the tissue-wide eQTL-GWAS method, the tissue with the most significant MR p-value was selected. In Figure S1, we show the proportion of genes mapped to a particular tissue category. The contribution of glandular-endocrine, neural central nervous system (CNS) and whole blood (eQTLGen) tissue categories were the highest (respective means of 15.3%, 12.8%, and 12.6% across the thirty traits; Table S4-5). Although each trait had genes mapped to nearly all tissues, a few distinctive patterns could be observed: cardiac muscle tissues contributed the most to atrial fibrillation (16.4%), vascular tissues the most to coronary artery disease (16.5%) followed by diastolic (11.1%) and systolic (9.9%) blood pressure, and the neural CNS contributed the most to schizophrenia (16.9%) and bipolar disease (16.6%).

**Figure 2:**
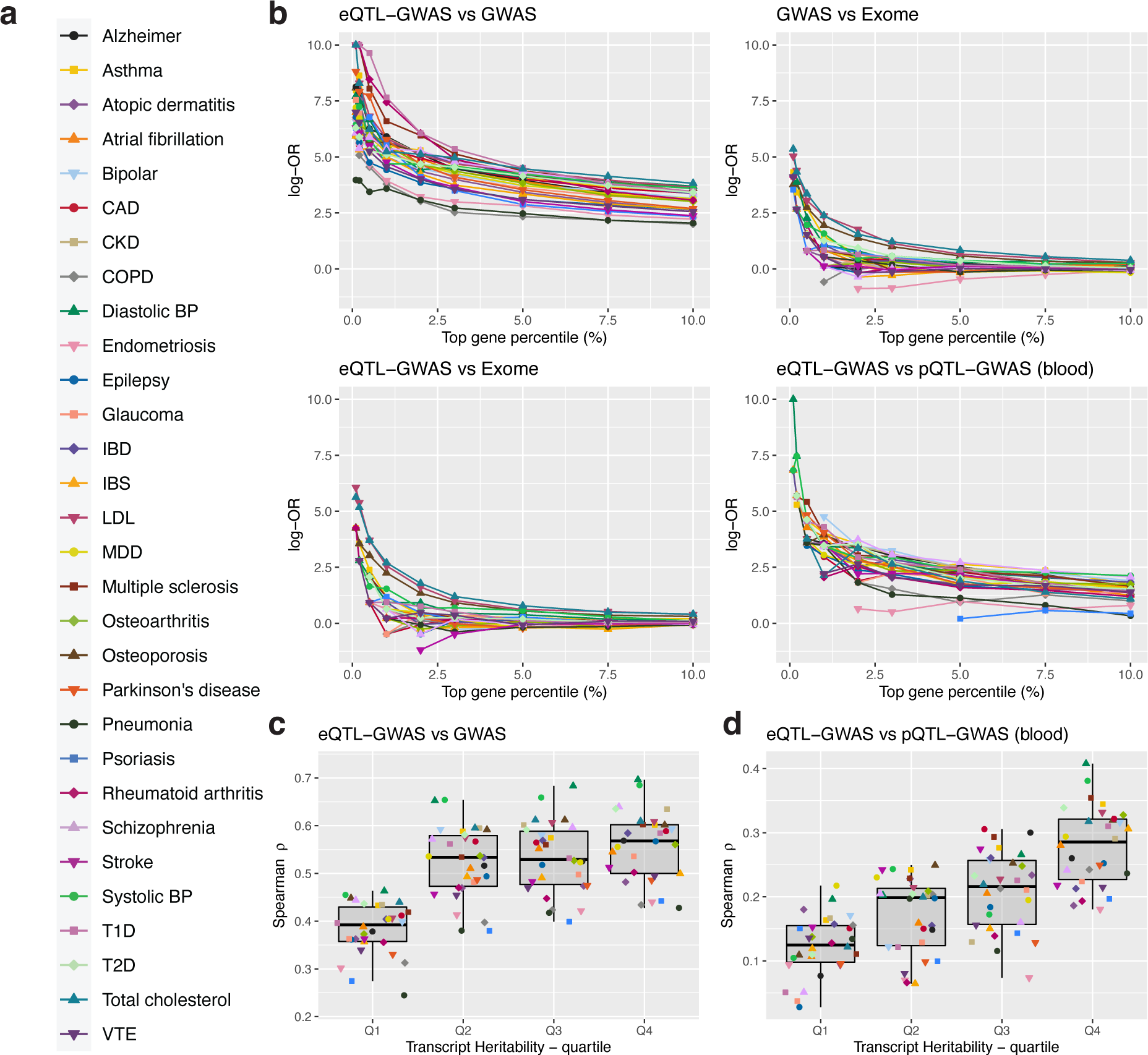
Evaluating the concordance of prioritized genes among methods. **a** Thirty different traits/drug indications were tested in this study. **b** The top prioritized genes between pairs of methods were compared at different thresholds for each trait (legend shown in **a**). The logarithm of odds ratios (log-OR) were calculated from Fisher’s exact tests. Log-ORs are only plotted for percentiles at which common genes between pairs of methods were found. Comparisons were conducted on the same background genes and same data origins (i.e., on UK Biobank GWASs for comparisons with the Exome method). Tissue-wide eQTL-GWAS gene prioritizations were considered for the comparison with the GWAS and Exome methods, and the blood-only eQTL-GWAS gene prioritization method for the comparison with the pQTL-GWAS method. **c** Spearman correlations calculated per trait between the gene significance (-log_10_(p-values)) from the GWAS and tissue-wide eQTL-GWAS methods. The analysis was conducted per trait on all common genes (*∼*16,100) stratified into quartiles according to transcript heritability (in *cis*). **d** Spearman correlations calculated per trait between gene Mendelian randomization (MR) causal effect sizes (*β*_MR_) from the blood eQTL-GWAS and pQTL-GWAS methods. The analysis was conducted on all common genes (*∼*1,370) stratified into quartiles according to the transcript heritability. In **c** and **d**, the boxplots bound the 25th, 50th (median, centre), and the 75th quantile. Whiskers range from minima (Q1 – 1.5 *·* IQR) to maxima (Q3 + 1.5 *·* IQR) with points corresponding to individual traits from **a**.

The concordance of prioritized genes among pairs of methods is summarized in Figure 2b. For each trait, we calculated Fisher’s exact tests between the top prioritized genes at thresholds ranging from the top 0.1%-10% (Methods). The overlap was the highest between the GWAS and eQTL-GWAS methods. At 1%, the median odds ratio (OR) was 212.2 which dropped to 51.0 and 22.1 at 5% and 10%, respectively. The concordance between GWAS and eQTL-GWAS prioritized genes was highest for transcripts with high heritabilities (Figure 2c; Methods). Similarly, the agreement between the eQTL-GWAS (whole blood) and pQTL-GWAS (blood plasma) increased with increasing transcript heritabilities (Figure 2d). Overall, the median ORs between these two methods was 8.5 and 4.6 at the top 5% and 10%, respectively. The overlap of prioritized genes was the lowest with the Exome method. The top 1% GWAS *vs* Exome and eQTL-GWAS *vs* Exome overlaps (based only on UKBB GWAS summary statistics), yielded median ORs of 1.7 and 1.9, respectively which dropped to 1.0 at 10% for both methods.

### Enrichment of prioritized genes for drug targets

Next, we assessed the extent to which prioritized genes overlapped with drug target genes. For each trait, we conducted enrichment analyses for the GWAS, eQTL-GWAS, pQTL-GWAS and Exome methods using our five definitions of drug target genes.

In Figure 3a, we show the resulting ORs for the DrugBank/DGIdb database combination. Across methods, genetic support for drug targets was the highest for LDL and total cholesterol (average ORs of 5.99 and 6.12, respectively). Lowest enrichment ratios were obtained for neuro-psychiatric traits (average OR of 1.56) and glaucoma (average OR of 1.14). The average OR across traits was 2.48, 2.68, 1.65, and 1.26 for the GWAS, eQTL-GWAS, Exome and pQTL-GWAS methods, respectively. We explored a range of top disease gene percentiles (0.1-5%) and the corresponding ORs are shown in Figure 3b. Restricting disease genes to the top 0.1% for all methods increased the average ORs without changing the method ranking with average ORs of 3.68, 4.02, 2.40, and 1.44 for the GWAS, eQTL-GWAS, Exome and pQTL-GWAS methods, respectively. We further analyzed whether identified drug targets were the same across methods and found that prioritized drug target genes were similar between GWAS and eQTL-GWAS methods (average Jaccard index of 0.39), less so between eQTL-GWAS and pQTL-GWAS methods (blood tissues; average Jaccard index of 0.15) and were very different from Exome identified targets (average Jaccard index of 0.06 between GWAS & Exome and eQTL-GWAS & Exome methods). Average AUC values across traits were of 53.4%, 51.9%, 50.5% and 49.9% for the GWAS, eQTL-GWAS, Exome and pQTL-GWAS methods (Figure 3c).

**Figure 3:**
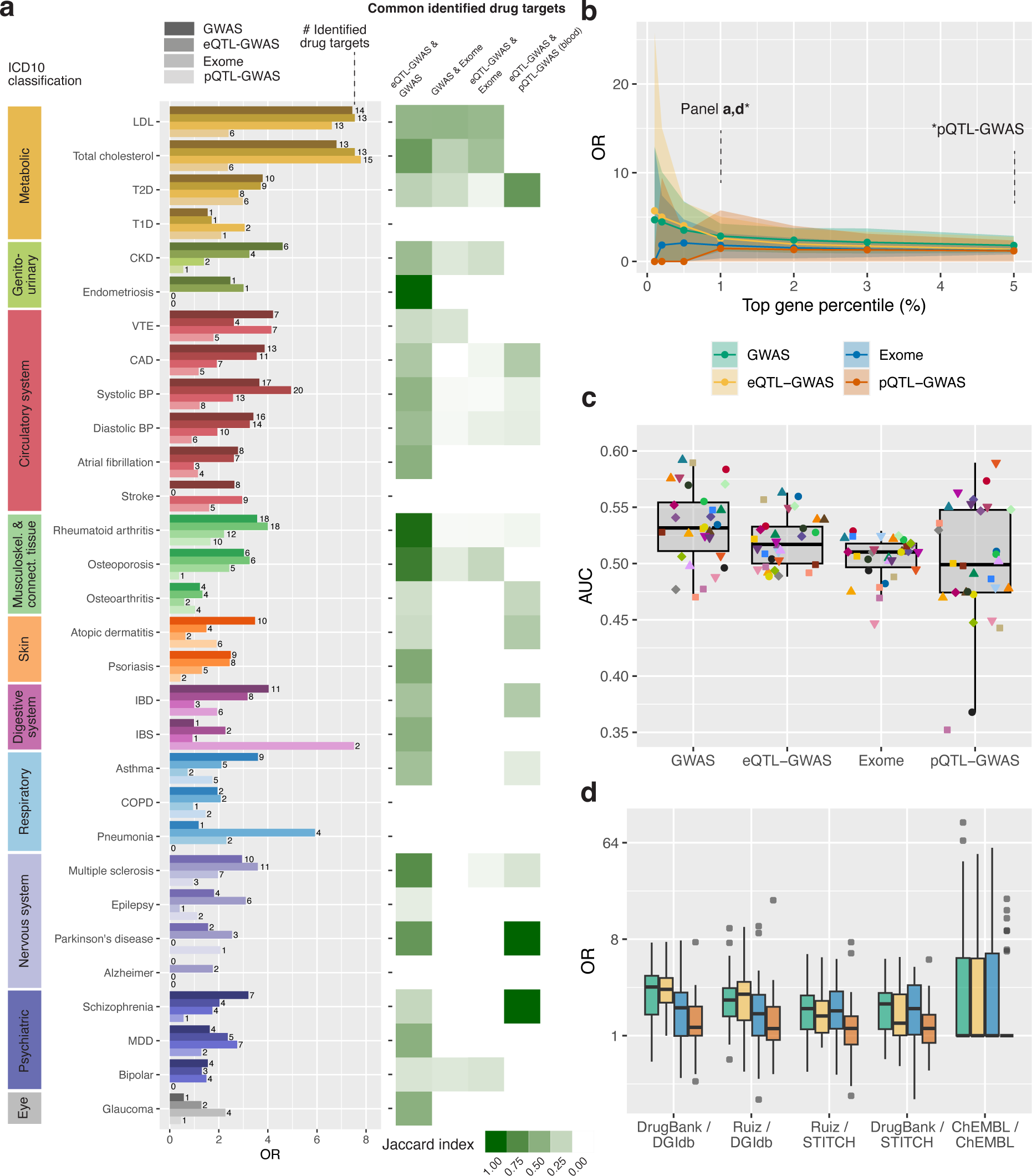
Enrichment for drug target genes. **a** Left: Barplot with odds ratios (ORs) calculated from Fisher’s exact tests between drug target genes and the top 1% (5% for pQTL-GWAS) prioritized genes for the four tested methods and thirty traits, classified according to trait category. Drug target genes were defined by DrugBank/DGIdb, and only drug target genes that could be tested by the respective method were considered. The number on the right of each bar indicates the number of identified drug target genes. Right: Overlap of identified drug target genes between pairs of methods quantified through the Jaccard index. The blood-only eQTL-GWAS gene prioritization method was used for the comparison with the pQTL-GWAS method. Plots using UKBB GWASs-only are shown in Figure S3. **b** ORs at different top prioritised gene percentiles for the four methods. The plotted dots correspond to the median OR across the thirty traits, and the shaded area bounds the 10% and 90% percentiles. **c** Boxplots showing the area under the receiver operating characteristic curve (AUC) values. AUC values were calculated for each trait as indicated by the points (legend in Figure 2a) and using the same background genes and drug target definitions as in **a**. **d** ORs calculated for the five drug target definitions and for all four methods (legend in **b**). The OR was set to 1 for traits with no identified drug target genes. In **b** and **d**, the boxplots bound the 25th, 50th (median, centre), and the 75th quantile. Whiskers range from minima (Q1 – 1.5 *·* IQR) to maxima (Q3 + 1.5 *·* IQR) with points outside representing potential outliers.

While the number of drugs reported per indication was similar across databases (average of 43.9, 41.8 and 40.4 for Ruiz *et al.*, ChEMBL and DrugBank, respectively), the average number of reported drug targets was much higher for Ruiz/STITCH (285), Ruiz/DGIdb (274.8), Drugbank/DGIdb (263.4) and Drugbank/STITCH (244.2) than for ChEMBL/ChEMBL (24.8; Table S7). We repeated drug target enrichment calculations for all drug database combinations (Figure 3d, Figure S2). The average OR for the GWAS/eQTL-GWAS methods were 2.48/2.68, 2.80/2.53, 2.18/2.12, 1.78/1.61 and 1.78/1.51 for Drugbank/DGIdb, ChEMBL/ChEMBL, Ruiz/DGIdb, Ruiz/STITCH and Drugbank/STITCH, respectively. Overall, the variability in ORs across traits was the highest in the ChEMBL database (Figure 3d, Figure S2), likely due to the low average number of reported drug targets which leads to very high ORs when drug targets figured among the prioritized genes (e.g. for LDL and total cholesterol), but for many traits drug target genes were not among prioritized genes (e.g. for type 1 diabetes, atopic dermatitis and inflammatory bowel disease).

Since enrichment results can differ widely across traits and reference databases, we calculated overall enrichment and AUC values across traits and drug databases, including sensitivity analyses on UKBB data only and common background genes (Table S9, Figure S4; Methods). The overall ORs were 2.17 (UKBB: 1.72), 2.04 (UKBB: 1.67), 1.81 and 1.31 (UKBB: 1.30) for the GWAS, eQTL-GWAS, Exome and pQTL-GWAS methods, respectively. There were no significant differences between these four methods in terms of enrichment OR (P_diff_ *>* 0.05 including in the sensitivity analyses). Overall AUCs were 54.3% (UKBB: 52.8%), 52.8% (UKBB: 51.4%), 51.7% and 51.3% (UKBB: 50.6%) for the GWAS, eQTL-GWAS, Exome and pQTL-GWAS methods, respectively. Judging by the AUC values, GWAS performed signifi-cantly better than eQTL-GWAS (P_diff_ = 3.1e-5) also when only considering testable eQTL genes (P_diff_ = 2.9e-4). Significantly higher AUC values were obtained for the GWAS compared to Exome on consortia data (P_diff_ = 2.2e-4) which was no longer the case on UKBB data (P_diff_ = 0.06). The difference between eQTL-GWAS and Exome was not significant on either dataset (P_diff_ of 0.12 and 0.77 on consortia and UKBB data, respectively). The number of testable genes was much lower for the pQTL-GWAS method (*∼*1,870 genes). With this set of background genes, GWAS still scored a higher overall AUC (55.1%, P_diff_ = 2.1e-3). No difference was observed between the pQTL-GWAS and tissue-wide or whole blood eQTL-GWAS methods (P_diff_ of 0.66 and 0.87, respectively).

### Examples of drug target prioritization ranks

In Figure 4, we highlight drug targets and their gene prioritization ranks for a few examples (complete list in Table S10). Major antihypercholesterolemic drug targets *PCSK9* (evolocumab, alirocumab), *HMGCR* (statins) and *NPC1L1* (ezetimibe) were top ranked by all methods (except for no pQTLs being available for *HMGCR* and *NPC1L1*; Figure 4a). HCN4, the target of antiarrhythmic drug dronedarone, was prioritized as a disease gene for atrial fibrillation only through the GWAS method. Although highly expressed in the atrial appendage and left ventricle of the heart, no eQTL was reported for this gene (Figure 4b). Several antiepileptic drugs target SCN1A which was highly prioritized by the GWAS and eQTL-GWAS methods with the strongest MR effect found in the nucleus accumbens (basal ganglia) of the brain (Figure 4c). Antiplatelet drug dipyrimadole used in the prevention and treatment of vascular diseases such as stroke and coronary artery disease is listed to target 23 genes of the *PDE* superfamily in ChEMBL. Of these, four (*PDE4D*, *PDE3A*, *PDE3B*, *PDE6B*) were ranked in the top 1% by the exome method for stroke (Figure 4d). None of the other methods prioritized any of these 23 genes. For coronary artery disease, another superfamily member (*PDE5A*) had a low ranking (*<* 2%) for the GWAS and QTL-GWAS methods, supported by solid GWAS and e/pQTL colocalisation (Figure 4e).

**Figure 4:**
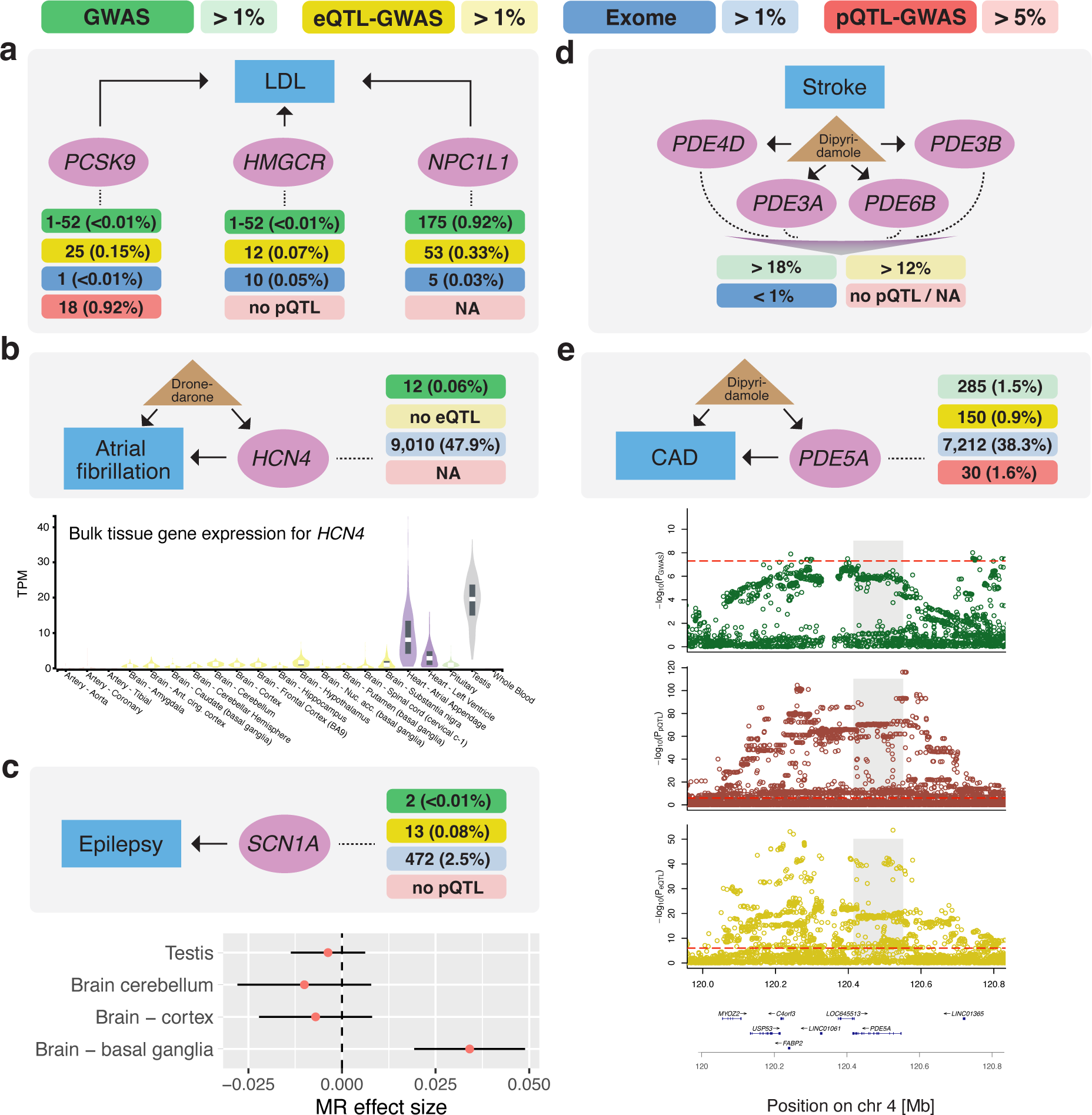
Examples illustrating drug target genes and their prioritization ranks. **a** Three drug target genes (*PCSK9* (evolocumab, alirocumab), *HMGCR* (statins) and *NPC1L1* (ezetimibe) shown in purple) for LDL cholesterol (blue box) and their prioritization ranks (top percentiles shown in parentheses) of each of the four methods (GWAS in green, eQTL-GWAS in yellow, Exome in blue and pQTL-GWAS in red). Genes that were not testable by a given method are reported as NA (no e/pQTL means that the gene was measured, but had no QTL) and a range of ranks (i.e., 1-52) indicates tied p-values. **b** Top plot shows the prioritization ranks of *HCN4*, the target of antiarrhythmic drug, dronedarone. Bottom plot shows the gene expression profile of *HCN4* across GTEx tissues (transcript per million - TPM) with “Testis”, “Heart – Atrial Appendage” and “Heart – Left Ventricle” dominating. **c** Top plot shows the prioritization ranks of *SCN1A* (Sodium Voltage-Gated Channel Alpha Subunit 1), a drug target gene of several antiepileptic drugs. Bottom plot shows Mendelian randomization (MR) effects (red dot) wih 95% CI (black bars) across tissues in which there was a significant eQTL. **d** Antiplatelet drug dipyrimadole and gene prioritization ranks of its multiple drug targets (a non-exhaustive selection) of the phosphodiesterase (*PDE*) superfamily. **e** Top plot shows the gene prioritization ranks of *PDE5A*, another reported target for dipyrimadole. Bottom plot shows the regional SNP associations (-log_10_(p-values)) with coronary artery disease (CAD; GWAS, green), PDE5A protein (pQTL, red) and *PDE5A* transcript (eQTL, yellow), respectively (red dashed lines indicate the significance thresholds of the respective SNP association, and grey shade marks the position of *PDE5A*). Bottom row illustrates the positions and strand direction of the genes in the locus.

### Heritability and polygenicity of drug target transcripts and proteins

It has been shown that drug target genes are more likely to have lower residual variance intolerance scores (RVIS), i.e., are less tolerant to change [1]. With the availability of large-scale eQTL and pQTL data, we tested whether drug target transcript or protein levels may be less impacted by genomic variations.

To this end, we compared *cis*-heritabilities and polygenicities of drug target genes versus non-drug target genes that were measured in the respective studies (i.e., also those with no reported e/pQTLs; Methods). We conducted the analysis per trait and for each of the five drug target gene definitions. Overall, we observed that drug target genes were less heritable and more polygenic than non-drug target genes pointing towards a negative selection [40] (Figure S5). This trend was less pronounced for tissue-wide than for whole blood eQTLs, and was the strongest for pQTLs. Across QTL studies and drug databases, the traits for which drug targets were significantly less heritable and more polygenic differed substantially. However, in no scenario drug target genes were more heritable than non-drug target genes on either the transcript or protein-level (at a nominally significant level). Considering drug target genes defined by DrugBank/DGIdb for the plasma protein QTLs as an example: targets for traits from the circulatory system were all shown to be more polygenic, and targets for psychiatric traits were all less heritable at a nominally significant level.

### Network diffusion

Finally, we assessed whether network diffusion can identify drug target genes for which there is no direct genetic evidence. Gene scores from prioritization methods defined the initial distribution ***p*_0_** of the diffusion process. This process is regulated by a restart parameter *r* where lower values result in a wider diffusion (i.e., genes can be prioritized even when distant from initial disease genes; Methods). The stationary distribution was calculated for six different restart parameters, ranging from no diffusion (r = 1) to complete diffusion (r = 0). Since the set of testable proteins (*∼*1,870) is enriched for drug target genes (two-sided binomial test: p-value = 1.3e-47 for DrugBank/DGIdb; complete results in Table S15; Methods) AUC values were artificially inflated upon projecting the gene scores onto the network and pQTL-GWAS results are hence not discussed.

Applying diffusion using the STRING network massively boosted the overlap between the diffused prioritized genes and the drug target genes (Figure 5a-b, Figure S6-7). At no diffusion, overall AUC values across the thirty traits were 54.3%, 52.8% and 51.7% for the GWAS, eQTL-GWAS and Exome methods, respectively, which increased to 68.9%, 67.7% and 66.9% at *r* = 0.6, and 73.5%, 72.9% and 72.3% at *r* = 0.4 (Figure 5a, Figure S6, Table S12). The same trend was observed when calculating enrichment scores for the top 1% genes with overall ORs of 4.63, 5.21 and 5.07 at *r* = 0.4 (Figure 5b, Figure S7, Table S12). On the other hand, improvements were modest when considering co-expression networks. At *r* = 0.6, overall AUC values increased to 54.9%, 54.7% and 53.5% in case of the CoXRNAseq network. Although small, the difference was significant compared to no diffusion (P_diff_ of 5.11e-3, 4.12e-14 and 4.83e-5, respectively). Overall ORs at *r* = 0.6 were 2.28, 2.04 and 1.91 which were not significantly different (P_diff_ *>* 0.05) compared to no diffusion. Likewise, in the FAVA network, overall AUC values at *r* = 0.6 were 55.9%, 54.2% and 53.6% (P_diff_ compared to no diffusion of 2.23e-5, 3.08e-3 and 7.3e-6), and ORs 2.38, 2.02 and 1.77 (P_diff_ *>* 0.05) for GWAS, eQTL-GWAS and Exome methods, respectively (Figure S6-7, Table S12-13).

**Figure 5:**
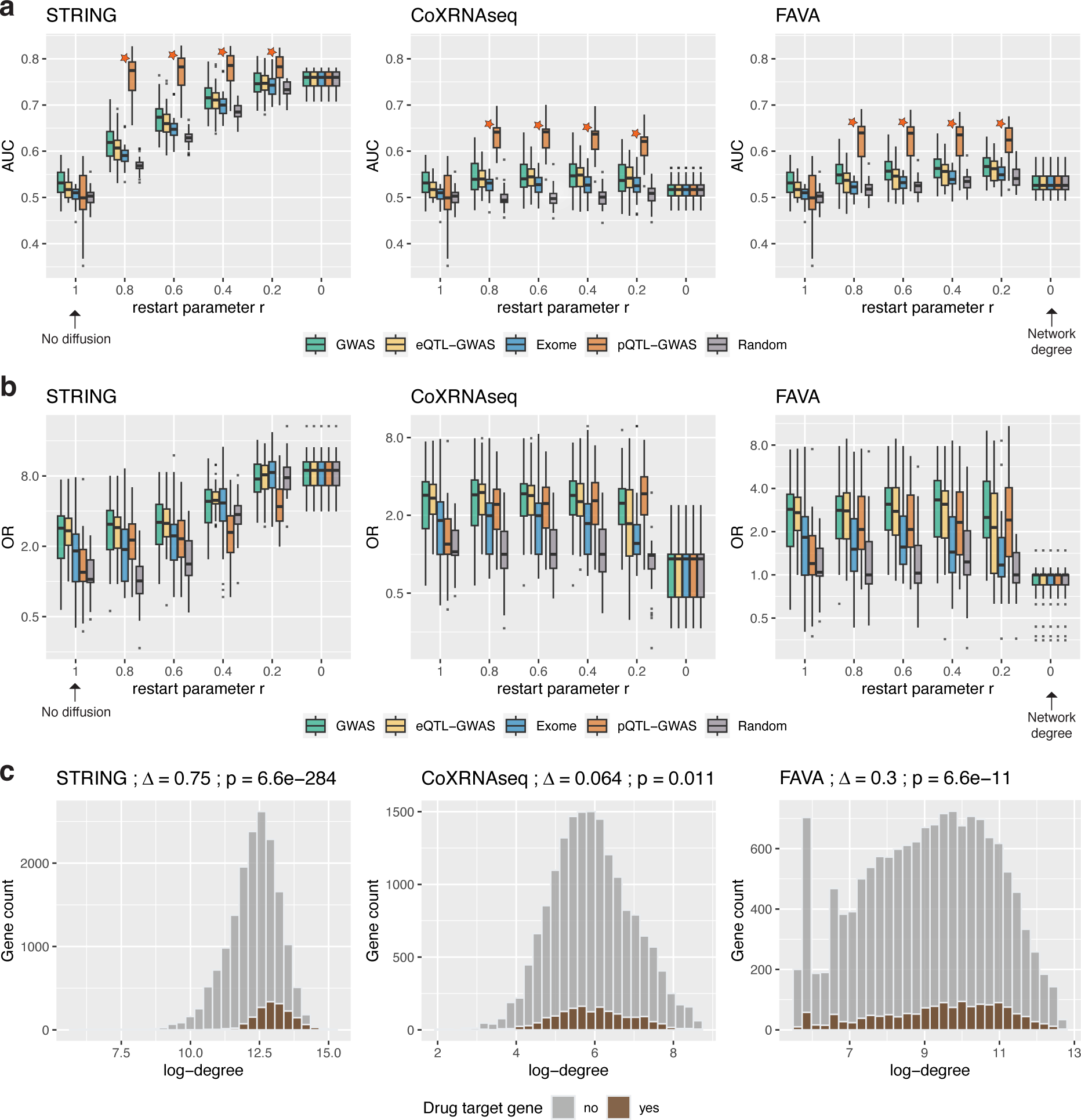
Effect of network diffusion to prioritize drug target genes. **a** Boxplots showing the area under the receiver operating characteristic curve (AUC) values for each network type (STRING, CoXRNAseq and FAVA) and method at different restart parameter values *r*. AUC values were calculated for each of the thirty traits, and drug target genes were defined by DrugBank/DGIdb. At an *r* value of 1 (no network diffusion), the analysis corresponds to the results in Figure 3b, and at an *r* value of 0, the gene prioritization rank is simply based on the degree of the network nodes. At *r <* 1, the background genes are the genes reported in the respective network. The star next to the pQTL-GWAS method signals that the set of testable genes for this method is enriched for drug target genes and therefore higher AUC values were obtained when adding background genes with zero-valued initial scores. **b** Odds ratios (ORs) between prioritized genes (top 1%) and drug target genes for each network type and method at different *r* values across the thirty traits (same drug target and background genes as in **a**). The OR was set to 1 for traits with no identified drug target genes. **c** Histograms showing the degree distribution of drug target genes and non-drug target genes in each network. The difference in log-degree (Δ) and the p-values from two-sided t-tests are shown in the titles. In **a** and **b**, the boxplots bound the 25th, 50th (median, centre), and the 75th quantile. Whiskers range from minima (Q1 – 1.5 *·* IQR) to maxima (Q3 + 1.5 *·* IQR) with points above or below representing potential outliers.

We further assessed which method’s AUC values benefited the most from network diffusion. To allow fair comparisons with the Exome methods, we used UKBB GWAS data for the GWAS and eQTL-GWAS methods. Across all diffusion parameters *r*, overall AUC values were significantly higher for GWAS than eQTL-GWAS in the STRING and FAVA network (P_diff_ *<* 4.45e-4), but not any different in the CoXRNAseq network (P_diff_ *>* 0.05). A nominally significant difference in favour of GWAS compared to Exome was only observed in the STRING network at *r* values of 0.4, 0.6 and 0.8 (P_diff_ of 0.0262, 7.36e-3 and 0.0146, respectively). No statistical differences were observed between the eQTL-GWAS and Exome method except for a nominally significant difference in favour of eQTL-GWAS at r = 0.2 in the CoXRNAseq network (P_diff_ = 0.0113).

When investigating the network connectivity, we observed that drug target genes were significantly more likely to be hub genes (Figure 5c, Figure S8). This observation was particularly strong in the STRING network (mean log-degree = 13.0 *vs* 12.3, P_diff_ = 6.6e-284 for DrugBank/DGIdb), but also present in the co-expression networks (Δ log-degree = 0.064, P_diff_ = 0.011 for CoXRNAseq; Δ log-degree = 0.3, P_diff_ = 6.6e-11 for FAVA). As a consequence, the network’s node degree was found to be a good predictor of drug targets, and the best performance was found for the network degree in STRING (overall AUC = 77.6%, overall OR = 8.71). Given this bias, we generated random initial disease gene scores and de-termined to which extent genetically-informed ***p*_0_** distributions performed better compared to random ***p*_0_** distributions. Although GWAS, eQTL-GWAS and Exome methods had significantly higher AUC values than random score distributions for any given *r* value in the STRING network (P_diff_ *<* 1.62e-7; Table S13), the performance of a mildly diffused (*r* = 0.8) random score (which is unaware of the target disease) per-formed significantly better than any disease gene prioritisation method without diffusion (P_diff_ of 4.18e-6, 3.58e-10 and 2.10e-12 compared to GWAS, eQTL-GWAs and Exome, respectively). In line with this observation, the network degree was still significantly better than gene prioritization methods at *r* = 0.2 (P_diff_ of 8.98e-6, 9.87e-13 and 1.89e-11 compared to GWAS, eQTL-GWAs and Exome, respectively).

### Examples of prioritized genes through network diffusion

In the following, we describe several examples for which drug targets figured among the top 1% genes only after network diffusion (complete list in Table S14). Amyloid-beta precursor protein (APP) targeted by the monoclonal antibody aducanumab in the treatment of Alzheimer’s disease was ranked 506 (top 2.7%) prior and 152 (top 0.8%) after diffusion on the STRING network (*r* = 0.6; Figure 6a) based on the eQTL-GWAS method. Prioritization was largely influenced by its interacting neighbour Apolipoprotein E (APOE) which was the top 5 ranked gene for Alzheimer’s disease. Tumor necrosis factor (TNF), a drug target in the treatment of inflammatory diseases such as psoriasis, was ranked 1,558 (top 8%; Exome-Psoriasis) prior and 182 (top 0.98%; *r* = 0.6) post-propagation in the STRING network (Figure 6b). While initially the drug target F2 (Coagulation Factor II, Thrombin) for VTE (venous thromboembolism) ranked only in the top 2% it moved up to the top 1% regardless of the network used for diffusion at *r* = 0.6 (top 0.9%, 0.6% and 0.7% for STRONG, CoXRNAseq and FAVA, respectively). In the STRING and CoXRNAseq networks, this boost could largely be attributed to the interacting fibrinogen genes (FGA, FGB and FGG) that ranked in the top 0.06% (Figure 6c).

**Figure 6:**
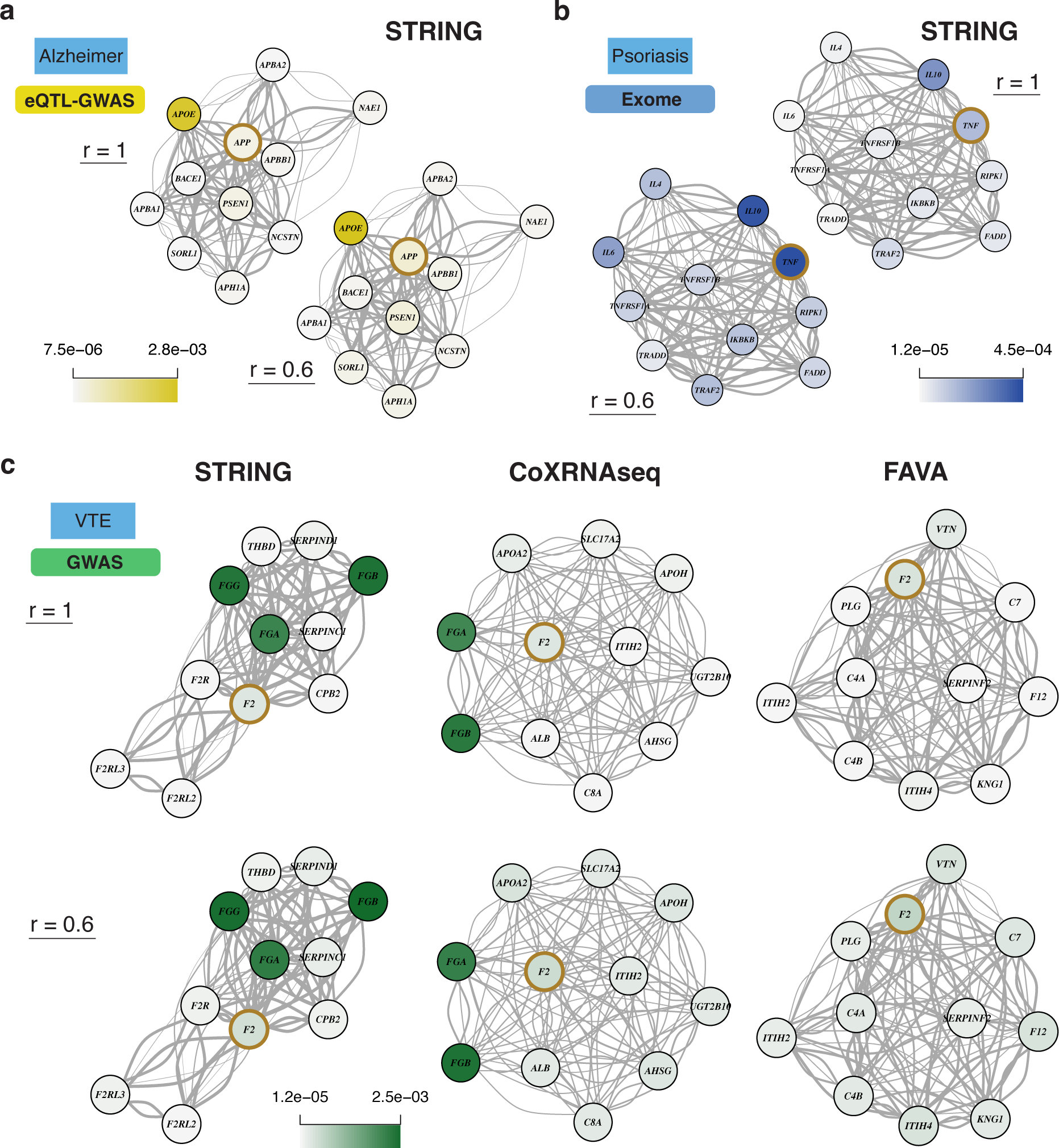
Examples illustrating network diffusion to prioritize drug target genes. **a** Top ten network neighbours of drug target APP (brown circle) and their prioritization values (i.e., normalized node probabilities) of the eQTL-GWAS method for Alzheimer’s disease are shown before (*r* = 1) and after diffusion (*r* = 0.6) on the STRING network. **b** Same representation as in **a** showing Exome prioritization values for psoriasis and tumor necrosis factor (TNF) drug target. **c** Top ten network neighbours of drug target F2 (Coagulation Factor II, Thrombin) in the STRING, CoXRNAseq and FAVA networks. GWAS prioritization values for venous thromboembolism (VTE) are shown before (*r* = 1) and after diffusion (*r* = 0.6) on each network. In each network example (**a**-**c**), the drug target gene was among the top 1% prioritized genes only after diffusion at *r* = 0.6.

## Discussion

We conducted a comprehensive benchmarking between different genetically informed approaches (GWAS, QTL-GWAS and Exome) combined with network diffusion to prioritize drug target genes. The strength of our analysis lies in the side-by-side comparison of gene prioritization methods that individually have proven to be successful in identifying drug targets. Recently, methods have emerged that combine multiple genetic predictors to derive an aggregate score often using machine-learning techniques [27, 41, 42]. These scores demonstrated high enrichment for drug targets but reveal little about underlying molecular mechanisms. Our aim was to disentangle the importance of the choice of the ground truth (i.e., drug target genes), the input data (such as molecular QTLs, WES) in combination with different molecular networks to highlight added benefits while also exposing weaknesses compared to using GWAS data alone.

Adjusting for differences in background genes and data origins, GWAS yielded higher AUC than eQTL- and pQTL-GWAS, but no significant difference was found with Exome. Genes prioritized by the Exome method were different from those identified by the GWAS and QTL-GWAS methods which was also reflected in the identified drug targets. While this could imply that rare- and common variant genetic architectures are complementary, differences could also be due to power issues. Possibly, with increased sample size the implicated genes will converge, but the extent to which they can be perturbed by regulatory *vs* rare coding variants might remain different. Considering ORs, we lacked statistical power to claim significant differences between methods since the number of drug targets among top 1% prioritized genes can be very low. Overall enrichment ORs for drug targets were 2.17, 2.04, 1.81 and 1.31 for the GWAS, eQTL-GWAS, Exome and pQTL-GWAS methods, respectively. Although ORs for the pQTL-GWAS method may seem lower, it should be noted that testable proteins (i.e., proteins with pQTLs) accounted for *∼*10% of GWAS testable genes. On the same background genes, ORs for the tissue-wide and blood-only eQTL-GWAS methods were 1.38 and 1.22, respectively. For the AUC metric, no significant difference between eQTL-GWAS and pQTL-GWAS was found. In the method comparisons, we considered multiple drug target gene definitions. The number of targets per drug drastically differed between ChEMBL and the DGIdb or STITCH databases due to differences in their construct. Drug target genes in the ChEMBL database are manually curated and should not contain false positives but it remains debatable whether to consider only primary or also secondary target genes. For instance, ChEMBL only lists HMGCR as a drug target for statins, whereas the DGIdb database also includes APOA5, APOB and APOE among others. For this reason, we considered different databases and present enrichment results for both broad and narrow drug target definitions, as well as aggregates.

Network diffusion was beneficial for prioritizing drug target genes with weaker genetic support. A remarkable increase in drug target identification was achieved when using the STRING network. However, this improvement may be due to a circularity in the data generation process leading to drug targets being much more likely to also be hub genes. Although genetically-informed methods performed better than random distributions, the STRING network node degrees resulted in the highest AUC values overall. Thus, care has to be taken when relying on literature-derived gene-gene interactions as aggressive diffusion will point to the same drug target genes, irrespective of the disease, and the intrinsic bias stemming from under- and over-studied genes may hinder the discovery of new drug targets. The improvements made with co-expression networks, which do not suffer from publication/curation biases, were minor in comparison. Although significant with the AUC metric, ORs were not significantly increased with a diffusion of *r* = 0.6 as compared to no diffusion for any of the methods.

Among the set of measured proteins (which itself was enriched for drug target genes), protein levels of drug target genes exhibited lower heritabilities and higher polygenicities than non-drug target genes. The same trend, but to a lesser extent, was observed on the transcript level. Additionally, drug target genes had stronger network connections than non-drug target genes, even in unbiased co-expression networks. Together, these observations suggest a tendency for drug targets to be evolutionary constrained and as a result are more protected from genomic perturbations.

Several limitations should be considered. First, we do not take into account directionality of therapeutic and genetic effects, i.e., whether the drug is an agonist or antagonist. Although found to be less performant than GWAS, QTL-GWAS methods have the advantage of specifying directionality, as opposed to gene scores from the GWAS approach which ignores SNP effect directions. Second, the used molecular QTL data sets cover only a small fraction of possible intermediate traits through which SNPs exert their disease-inducing effects [43]. Third, we only focus on common genetic variants when associating transcript and protein levels. With the advent of coupled rare variant-protein level data, either from populations enriched for rare variants or sequencing data [14, 44], more powerful QTL-GWAS methods are likely to emerge that combine mechanistic insights gained from QTL approaches while capturing rare variant associations previously missed. Fourth, drug target data are sparse which limits the statistical power in benchmarking analyses. Given the required resources to test a drug target in clinical settings, focusing on top ranking genes is of most interest. This scenario is best described with a threshold that defines highly prioritized genes for enrichment analyses. However, ROC curves that quantify the performance at all prioritization thresholds (i.e. use all data at hand) were better powered to detect subtle differences between methods. Finally, our analysis compares methods using historical drug discovery data as the ground truth. This data is highly biased with G-protein-coupled receptors being targets of a third of FDA-approved drugs [45]. Many other genes may be effective targets, but have never been tested in clinical trials. Thus, our results may not reflect how well the tested genetic approaches uncover true disease genes, but rather how well they identify targets that were historically prioritized in drug development processes. Since the emergence of robust GWAS, more and more clinical trials are motivated by genetically informed targets. Thus, drug tar-get databases will tend to overlap better with GWAS-inspired genes, leading to artificially higher overlap.

To conclude, we systematically evaluated major gene prioritization approaches for their ability to identify approved drug target genes. Our analyses highlight the power of harnessing multiple data sources by capitalizing on QTLs for mechanistic insights, sequencing data for rare variant associations, GWAS when molecular QTL signals are missing and network propagation to leverage gene-gene interactions.

## Methods

### GWAS data

We used the largest (to-date), publicly available GWAS summary statistics for each analyzed condi-tion (Table S1). GWAS data came mostly from consortia specific to the respective disease, and were often a meta-analysis comprising the UKBB. Twenty-four out of the 30 conditions were case/control studies, the remaining 6 being continuous traits: diastolic and systolic blood pressure (DBP and SBP, re-spectively [46]), low-density lipoprotein and total cholesterol (LDL and TC, respectively [47]), estimated glomerular filtration rate (eGFR [48]) and heel bone mineral density ([47]) proxying chronic kidney dis-ease (CKD) and osteoporosis, respectively. For four traits with low case count in the UK Biobank (*<* 20,000; chronic obstructive pulmonary disease (COPD), endometriosis, pneumonia and psoriasis) and no large-scale GWAS meta-analysis available, we performed a meta-analysis between the UK Biobank [47] and FinnGen [49] using METAL [50].

### GWAS gene scores

We used PascalX [9, 51] to compute gene scores based on GWAS summary statistics. The software takes as input GWAS p-values, gene annotations and LD structure. SNPs are assigned to genes and their squared z-scores are summed. This sum, under the null, was shown to follow a weighted chi-square distribution with weights being defined by the local LD structure from which gene p-values can be derived [9]. We applied PascalX with default parameters (gene *±* 50 kB) on protein-coding genes using the Ensembl identifiers and annotations (Ensembl GRCh37.p13 version) and the UK10K reference panel [52]. Across traits, *∼*19,150 protein-coding genes could be tested which were ranked by their PascalX p-value.

### Molecular QTL-GWAS gene scores

We integrated molecular quantitative trait loci (QTL) and GWAS summary statistics using Mendelian randomization (MR) implemented in the smr-ivw software [53, 22]. The exposure (transcript or protein levels) and outcome disease were instrumented with independent genetic variants, also called instrumental variables (IVs; *r*^2^ *<* 0.01) and used to calculate putative causal effect estimates of the exposure on the outcome (*β*_MR_). IVs were required to be strongly associated to the exposure (P_QTL_ *<* 1e-6) and had to pass the Steiger filter ensuring no significantly stronger effect on the outcome than on the exposure [54]. We used expression QTLs (eQTLs) from the eQTLGen consortium [13] (whole blood; *n* = 31,684) and tissue-specific QTLs from the GTEx v8 release [55] (European ancestry; *n* = 65-573 for 48 tissue types; Table S4) to estimate causal transcript-trait effects. In the eQTLGen dataset there were *∼*12,550 protein-coding genes with at least 1 IV which increased to *∼*16,250 when integrating the GTEx dataset. MR results from both datasets (whole blood from eQTLGen and 49 tissues from GTEx) were aggregated by considering the MR causal effect with the lowest p-value across tissues (Table S4-5). Protein QTLs (pQTLs) from the deCODE study [14] (whole blood; *n* = 35,559) were used to estimate protein-trait causal effects with *∼*1,870 proteins having at least 1 IV. Prior to the analysis, e/pQTL and GWAS data were harmonized, palindromic SNPs were removed as well as SNPs with an allele frequency difference *>* 0.05 between datasets. All transcripts and proteins were mapped to Ensembl identifiers as provided by eQTLGen, GTEx and deCODE.

### Exome gene scores

We used gene burden test results computed on WES data from the UK Biobank [25]. We extracted gene-trait associations based on putative loss of function (pLOF) and deleterious missense variants with MAF *<*1% (M3.1 nomenclature in original publication) with phenotypes matching the investigated conditions as indicated in Table S2. Associations were provided for *∼*18,800 genes which were ranked by the association p-value and retrieved by the provided Ensembl identifier.

### Concordance of gene scoring methods

We tested whether prioritized genes were similar or dissimilar between pairs of methods. First, only genes (based on Ensembl identifiers) that were common between the two tested methods were selected into the gene background. Then, prioritized genes were defined at different top percentile cut-offs (0.1%, 0.2%, 0.5%, 1%, 2%, 3%, 5%, 7.5%, 10%). The enrichment of prioritized genes between methods was quantified by a Fisher’s exact test using common genes as background genes. When calculating median ORs, ORs of traits for which no prioritized genes overlapped at a given percentile were set to 1.

### Drug target genes

We extracted drug target genes from public resources by combining drug-indication and drug-target links from various databases. A given disease/indication was linked to a drug if the drug was indicated to be prescribed for the selected indication and subsequently, the target genes of these drugs were extracted. For drug-indication pairs we consulted DrugBank, Ruiz *et al.* and ChEMBL:

- DrugBank 5.0 [35] (download: May 2022): DrugBank indications are manually curated from drug labels and underwent an expert review process. Drug indications have their own DrugBank condition numbers and drugs their DrugBank identifiers.
- Ruiz *et al.* [36]: A drug-disease dataset was created by querying multiple sources such as the Drug Repurposing Database, the Drug Repurposing Hub, and the Drug Indication Database and extracting information from drug labels, DrugBank and the American Association of Clinical Trials Database. Drug–disease pairs were filtered for FDA-approved treatment relationships. This dataset uses NLM UMLS CUIDS identifiers (National Library of Medicine -Unified Medical Language System Controlled Unique Identifier) for diseases and DrugBank identifiers for drugs.
- ChEMBL [37] (download: May 2022): ChEMBL drug indications are extracted from multiple sources including DailyMed package inserts, Anatomical Therapeutic Chemical (ATC) classification and ClinicalTrials.gov. Mapping of disease terms to Medical Subject Headings (MeSH) vocab-ulary and the Experimental Factor Ontology (EFO) is done through a combination of text-mining, automated mapping and manual curation/validation. Drugs are reported with ChEMBL identifiers.

The mapping of GWAS traits to the drug indication identifiers of the respective database is shown in Table S6.

Drug target genes were extracted from the DGIdb, STITCH and ChEMBL databases:

- Drug Gene Interaction database (DGIdb) 4.0 [38] (release: January 2021): Aggregated drug-gene interactions from multiple sources including DrugBank, Drug Target Commons, the Therapeutic Target Database and Guide to Pharmacology. Genes were matched to Ensembl identifiers using the provided gene vocabulary file. Drugs were reported through DrugBank or ChEMBL identifiers, and mapping from ChEMBL to DrugBank identifiers was done with UniChem [56], using PubChem IDs as intermediates.
- Search Tool for Interacting CHemicals (STITCH) 5.0 [39]: Aggregated drug-protein interaction data from high-throughput experiments data, manually curated datasets and prediction methods. Only high confidence drug-protein relationships (confidence score *≥* 700) of the type “inhibition” and “activation” were considered. STITCH uses PubChem Chemical Identifiers (CID) for drugs and mapping to DrugBank IDs was done through the *chemical sources* file provided by STITCH. Protein Ensembl identifiers were mapped to gene Ensembl identifiers using biomaRt (GRCh37, v2.50.3)[57].
- ChEMBL [37] (download: May 2022): ChEMBL provides drug targets which have been manually curated from literature. Drug targets are identified by ChEMBL IDs with mapping to UniProt Accessions provided by ChEMBL. UniProt identifiers were then mapped to gene Ensembl identifiers through the UniProt REST API [58].

In this analysis we considered drug target genes resulting from the following combinations: Drug-Bank/DGIdb, DrugBank/STITCH, Ruiz/DGIdb, Ruiz/STITCH, and ChEMBL/ChEMBL. The number of drugs and drug target genes per indication is shown in Table S7.

### Drug target enrichment and AUC calculations

Enrichment for drug target genes was calculated through two-sided Fisher’s exact tests. A contingency table was constructed based on testable genes (i.e., background genes), with genes categorized into prioritized (top 1% or 5% for the pQTL-GWAS) and drug target genes. In rare instances (i.e., pQTL-GWAS background genes and ChEMBL/ChEMBL drug targets) where diagonal values were 0, these were changed to 1. If no prioritized gene coincided with a drug target gene, the resulting OR was set to 1 (for visualization purposes this was not done in barplots where each trait was shown individually). AUC values and standard errors were calculated using the R package pROC v1.15.3 [59].

Log-OR and AUC values (both are denoted *b_i_* herein) were aggregated across traits and drug databases (*m* = 30 *·* 5 = 150 observations per method) as follows:

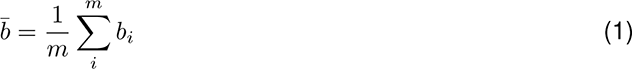

with corresponding variance:

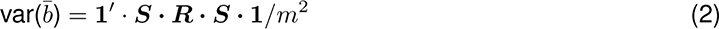

where ***S*** is a diagonal matrix of size *m*x*m* containing standard errors of *b_i_* and ***R*** is the correlation matrix between drug databases and traits. This matrix was derived from the Kronecker product of the drug database correlation matrix and phenotypic trait correlation matrix (Table S8). The drug database correlation matrix was derived on the gene level (i.e., 1 if the gene was a drug target for any of the 30 traits, 0 if not) and the phenotypic trait correlation on individual-level data from the UKBB (codes in Table S2). 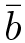 was referred to as the overall AUC/ log-OR (overall OR after an exponential transformation).

To calculate the statistical difference of 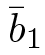 and 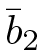 for method 1 and 2, respectively, we derived the variance of the difference as follows:

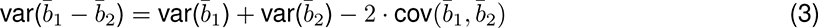

with cov(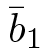, 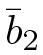) *≈ r ×* (**1***^′^ · **S*****_1_** · ***R*** · ***S*_2_** · **1***/m*^2^), where *r* is the empirical correlation between ***b***_1_ and ***b***_2_. From the resulting z-score, a two-sided p-value was calculated.

### Transcript and protein level heritabilities and polygenicity

Transcript and protein level heritabilities were estimated from QTL effects. Standardized QTL effects 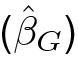 were approximated by dividing SNP z-scores by the square root of the sample size. The corre-sponding variance (var 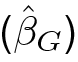) equals the inverse of the sample size. The *cis* heritability 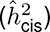 could then be estimated by summing up effects of independent (*r*^2^ *<* 0.01) and significant (p-value *<* 1e-6) QTLs in proximity of the transcript/protein (*±* 500 kB) while taking into account their variance:

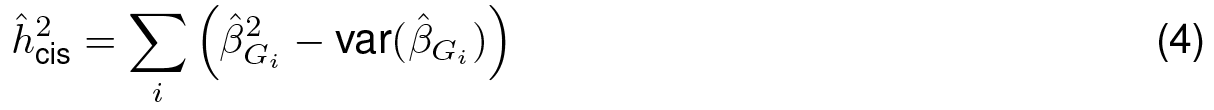

Protein heritabilities were based on the deCODE plasma protein dataset [14] and transcript heritabilities for whole blood on the eQTLGen dataset [13]. Tissue-wide transcript heritabilities were based on eQTL effects from the tissue in which the MR effect was the most significant [55].

To calculate the difference in heritabilities and polygenicities between drug target and non-drug target genes, we considered all transcripts and proteins measured in the respective study which were classified accordingly. Per trait, the difference in heritability was then calculated through a two-sided t-test. For the polygenicity analysis, we considered the number of independent (*r*^2^ *<* 0.01) and significant (p-value *<* 1e-6) QTLs associated to a transcript/protein. Note that this proxy for polygenicity is biased, as for less heritable omics entities we have less power to detect the independent signals, and may look less polygenic. We then conducted two-sided Wilcoxon tests to determine the difference in number of instrumental variables (IVs). Heritability and polygenicity tests were only performed for traits with at least three drug targets within the respective set of measured transcripts/proteins. Note that in case a transcript or protein was reported as measured in the study, but had no associated QTL, its heritability and number of IVs were set to zero.

### Networks

To calculate network diffusion scores, we used the following three networks:

- Search Tool for Retrieval of Interacting Genes/Proteins (STRING) v11 [32]: The protein-protein (PPI) interaction network results from predictions based on genomic context information, co-expression, text-mining, experimental biochemical/genetic data and curated databases (curated pathways and protein-complex knowledge). Protein Ensembl identifiers were mapped to gene Ensembl identifiers using biomaRt (GRCh37, v2.50.3)[57]. We use interaction confidence scores as edge weights.
- CoXRNAseq [33]: This network was constructed by first performing a principal component analysis on the gene co-expression correlation matrix of 31,499 RNA-seq samples. Reliable principal components were retained from which the final network was constructed via Pearson correlations. We filtered pairwise interactions to only retain those with z-scores above 4. Genes were reported with Ensembl identifiers and z-scores were used as edge weights.
- Functional Associations using Variational Autoencoders (FAVA) [34]: This network is based on single cell RNA-seq read-count data from the Human Protein Atlas and proteomics data from the PRoteomics IDEntifications (PRIDE) database. First, the high-dimensional expression data was reduced into a latent space using variational autoencoders. From this latent space, the network was derived via pairwise Pearson correlations. Each reported interaction has a score which we use as edge weight (final network reports interactions with scores above 0.15). Protein Ensembl identifiers were mapped to gene Ensembl identifiers using biomaRt.

A summary of network properties is given in Table S11. In all analyses, we use weighted networks, and we refer to weighted node degrees (i.e., sum of edge weights linking the node of interest to adjacent nodes) as node degrees.

### Network diffusion

We calculated network diffusion scores based on Markov random walks. Starting from an initial node distribution ***p*_0_**, a stationary distribution is calculated based on network connectivity. This diffusion process depends on a restart parameter *r* which determines how often the random walker returns to the initial values. Analytically, the stationary distribution (***p_∞_***) is given by:

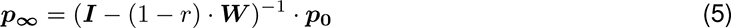

where ***W*** is the column-normalized weighted adjacency matrix and ***I*** the identity matrix of the same dimension as ***W*** [60]. The initial node distribution ***p*_0_** was determined by the squared z-scores derived from the gene p-values (normalized to sum up to 1). Genes that could not be tested by a given method had their initial value set to 0. Additionally, we tested the performance of network diffusion on random initial distributions ***p*_0_**. For each trait, a random distribution was generated which all were different, but consistent across analyses. Resulting network diffusion scores ***p_∞_*** were ranked for AUC calculations, and the top 1% scored genes were used in the enrichment analyses.

Network manipulations, visualization and degree calculations were performed with the R igraph package v1.3.5 [61].

### Enrichment of available proteins for drug targets

We conducted binomial tests to verify whether the set of testable (i.e., at least 1 pQTL) and measured proteins (*∼*1,870 and *∼*4,450, respectively) were enriched for drug target genes. We performed the analysis on each of the five drug target definitions and proceeded as follows: 1) we extracted the number of testable/measured proteins that are drug targets (“number of successes”), 2) considering all protein-coding autosomal genes (19,430), we extracted those that are drug targets (“number of trials”), 3) we determined the proportion of testable/measured proteins among all protein-coding genes (“expected probability of success”). From these numbers, we conducted two-sided exact binomial tests (Table S15).

## Data availability

Whole blood expression QTLs are from the eQTLGen eQTL meta-analysis and are available at https://www.eqtlgen.org/cis-eqtls.html. Tissue-wide expression QTLs are from the GTEx project and are available at https://gtexportal.org/home/datasets. Plasma protein QTLs are from the deCODE study and are available at https://www.decode.com/summarydata/. Summary statistics from whole exome gene burden tests are available in the GWAS Catalog (accession IDs are in Table S2). Genetic and phenotypic data from the UK Biobank Resource are available to approved researchers. GWAS summary statistics from the UK Biobank are available at http://www.nealelab.is/uk-biobank and https://pan.ukbb.broadinstitute.org. GWAS summary statistics from FinnGen are available at https://www.finngen.fi/en/access_results. GWAS summary statistics for multiple sclerosis (MS) are available by application from https://imsgc.net/?page_id=31. Full list of GWAS summary statistics used in this study is in Table S1-3, all of which are publicly available. UK10K individual-level data are available upon request (https://www.uk10k.org/data_access.html).

## Code availability

GWAS calculations were performed with REGENIE which is available at https://github.com/rgcgithub/ regenie. GWAS meta-analyses were performed with METAL which is available at https://github.com/statgen/METAL. Gene scoring was performed with the PascalX software which is available at https://github.com/BergmannLab/PascalX. QTL Mendelian randomization analyses were performed with the SMR-IVW software which is available at https://github.com/masadler/smrivw.

## Competing interests

The authors declare that they have no competing interests.

## Authors’ contributions

MCS and ZK conceived and designed the study. MCS performed statistical analyses. PD provided guidance on statistical analyses. ZK supervised all statistical analyses. CA contributed to the collection and interpretation of pharmacological and biological data. All the authors contributed by providing advice on interpretation of results. MCS and ZK drafted the manuscript. All authors read, approved, and provided feedback on the final manuscript.

## Acknowledgements

This work was supported by the Swiss National Science Foundation (310030 189147) to Z.K. This research has been conducted using the UK Biobank Resource under Application Number 16389. LD was calculated based on the UK10K data resource (EGAD00001000740, EGAD00001000741). Computations we performed on the JURA cluster of the University of Lausanne. We also would like to acknowledge the participants and investigators of the UK Biobank and FinnGen study. We thank Daniel Krefl for his help and support in implementing the PascalX software and Liza Darrous for critical reading of the draft.

## Notes

### Competing Interest Statement

The authors have declared no competing interest.

### Author Declarations

Ethics committee/IRB of UK Biobank gave ethical approval for this work.

## References

[1] Matthew R Nelson, Hannah Tipney, Jeffery L Painter, Judong Shen, Paola Nicoletti, Yufeng Shen, Aris Floratos, Pak Chung Sham, Mulin Jun Li, Junwen Wang, et al. The support of human genetic evidence for approved drug indications. Nature genetics, 47(8):856–860, 2015.

[2] Emily A King, J Wade Davis, and Jacob F Degner. Are drug targets with genetic support twice as likely to be approved? revised estimates of the impact of genetic support for drug mechanisms on the probability of drug approval. PLoS genetics, 15(12):e1008489, 2019.

[3] Stacey L Edwards, Jonathan Beesley, Juliet D French, and Alison M Dunning. Beyond gwass: illuminating the dark road from association to function. The American Journal of Human Genetics, 93(5):779–797, 2013.

[4] Chen Cao and John Moult. Gwas and drug targets. BMC genomics, 15(4):1–14, 2014.

[5] Vivian Tam, Nikunj Patel, Michelle Turcotte, Yohan Bossé, Guillaume Paré, and David Meyre. Bene-fits and limitations of genome-wide association studies. Nature Reviews Genetics, 20(8):467–484, 2019.

[6] Steven Gazal, Omer Weissbrod, Farhad Hormozdiari, Kushal K Dey, Joseph Nasser, Karthik A Jagadeesh, Daniel J Weiner, Huwenbo Shi, Charles P Fulco, Luke J O’Connor, et al. Combining snp-to-gene linking strategies to identify disease genes and assess disease omnigenicity. Nature Genetics, pages 1–10, 2022.

[7] Jimmy Z Liu, Allan F Mcrae, Dale R Nyholt, Sarah E Medland, Naomi R Wray, Kevin M Brown, Nicholas K Hayward, Grant W Montgomery, Peter M Visscher, Nicholas G Martin, et al. A versatile gene-based test for genome-wide association studies. The American Journal of Human Genetics, 87(1):139–145, 2010.

[8] Christiaan A de Leeuw, Joris M Mooij, Tom Heskes, and Danielle Posthuma. Magma: generalized gene-set analysis of gwas data. PLoS computational biology, 11(4):e1004219, 2015.

[9] David Lamparter, Daniel Marbach, Rico Rueedi, Zoltán Kutalik, and Sven Bergmann. Fast and rigorous computation of gene and pathway scores from snp-based summary statistics. PLoS computational biology, 12(1):e1004714, 2016.

[10] John Lonsdale, Jeffrey Thomas, Mike Salvatore, Rebecca Phillips, Edmund Lo, Saboor Shad, Richard Hasz, Gary Walters, Fernando Garcia, Nancy Young, et al. The genotype-tissue expression (gtex) project. Nature genetics, 45(6):580–585, 2013.

[11] Benjamin B Sun, Joseph C Maranville, James E Peters, David Stacey, James R Staley, James Blackshaw, Stephen Burgess, Tao Jiang, Ellie Paige, Praveen Surendran, et al. Genomic atlas of the human plasma proteome. Nature, 558(7708):73–79, 2018.

[12] Lasse Folkersen, Stefan Gustafsson, Qin Wang, Daniel Hvidberg Hansen, Å sa K Hedman, Andrew Schork, Karen Page, Daria V Zhernakova, Yang Wu, James Peters, et al. Genomic and drug target evaluation of 90 cardiovascular proteins in 30,931 individuals. Nature metabolism, 2(10):1135–1148, 2020.

[13] Urmo Võsa, Annique Claringbould, Harm-Jan Westra, Marc Jan Bonder, Patrick Deelen, Biao Zeng, Holger Kirsten, Ashis Saha, Roman Kreuzhuber, Harm Brugge, et al. Large-scale cis-and trans-eqtl analyses identify thousands of genetic loci and polygenic scores that regulate blood gene expression. Nature genetics, pages 1–11, 2021.

[14] Egil Ferkingstad, Patrick Sulem, Bjarni A Atlason, Gardar Sveinbjornsson, Magnus I Magnusson, Edda L Styrmisdottir, Kristbjorg Gunnarsdottir, Agnar Helgason, Asmundur Oddsson, Bjarni V Hall-dorsson, et al. Large-scale integration of the plasma proteome with genetics and disease. Nature Genetics, 53(12):1712–1721, 2021.

[15] Benjamin B Sun, Joshua Chiou, Matthew Traylor, Christian Benner, Yi-Hsiang Hsu, Tom G Richardson, Praveen Surendran, Anubha Mahajan, Chloe Robins, Steven G Vasquez-Grinnell, et al. Genetic regulation of the human plasma proteome in 54,306 uk biobank participants. BioRxiv, 2022.

[16] Claudia Giambartolomei, Damjan Vukcevic, Eric E Schadt, Lude Franke, Aroon D Hingorani, Chris Wallace, and Vincent Plagnol. Bayesian test for colocalisation between pairs of genetic association studies using summary statistics. PLoS Genet, 10(5):e1004383, 2014.

[17] Farhad Hormozdiari, Martijn Van De Bunt, Ayellet V Segre, Xiao Li, Jong Wha J Joo, Michael Bilow, Jae Hoon Sul, Sriram Sankararaman, Bogdan Pasaniuc, and Eleazar Eskin. Colocalization of gwas and eqtl signals detects target genes. The American Journal of Human Genetics, 99(6):1245–1260, 2016.

[18] Alexander Gusev, Arthur Ko, Huwenbo Shi, Gaurav Bhatia, Wonil Chung, Brenda WJH Penninx, Rick Jansen, Eco JC De Geus, Dorret I Boomsma, Fred A Wright, et al. Integrative approaches for large-scale transcriptome-wide association studies. Nature genetics, 48(3):245–252, 2016.

[19] Alvaro N Barbeira, Scott P Dickinson, Rodrigo Bonazzola, Jiamao Zheng, Heather E Wheeler, Jason M Torres, Eric S Torstenson, Kaanan P Shah, Tzintzuni Garcia, Todd L Edwards, et al. Exploring the phenotypic consequences of tissue specific gene expression variation inferred from gwas summary statistics. Nature communications, 9(1):1–20, 2018.

[20] Yang Wu, Jian Zeng, Futao Zhang, Zhihong Zhu, Ting Qi, Zhili Zheng, Luke R Lloyd-Jones, Riccardo E Marioni, Nicholas G Martin, Grant W Montgomery, et al. Integrative analysis of omics summary data reveals putative mechanisms underlying complex traits. Nature communications, 9(1):1–14, 2018.

[21] Eleonora Porcu, Sina Rüeger, Kaido Lepik, Federico A Santoni, Alexandre Reymond, and Zoltán Kutalik. Mendelian randomization integrating gwas and eqtl data reveals genetic determinants of complex and clinical traits. Nature communications, 10(1):1–12, 2019.

[22] Marie C Sadler, Chiara Auwerx, Kaido Lepik, Eleonora Porcu, and Zoltán Kutalik. Quantifying the role of transcript levels in mediating dna methylation effects on complex traits and diseases. Nature Communications, 13(1):1–14, 2022.

[23] Elizabeth T Cirulli, Simon White, Robert W Read, Gai Elhanan, William J Metcalf, Francisco Tanudjaja, Donna M Fath, Efren Sandoval, Magnus Isaksson, Karen A Schlauch, et al. Genome-wide rare variant analysis for thousands of phenotypes in over 70,000 exomes from two cohorts. Nature communications, 11(1):1–10, 2020.

[24] Jack A Kosmicki, Julie E Horowitz, Nilanjana Banerjee, Rouel Lanche, Anthony Marcketta, Evan Maxwell, Xiaodong Bai, Dylan Sun, Joshua D Backman, Deepika Sharma, et al. Pan-ancestry exome-wide association analyses of covid-19 outcomes in 586,157 individuals. The American Journal of Human Genetics, 108(7):1350–1355, 2021.

[25] Joshua D Backman, Alexander H Li, Anthony Marcketta, Dylan Sun, Joelle Mbatchou, Michael D Kessler, Christian Benner, Daren Liu, Adam E Locke, Suganthi Balasubramanian, et al. Exome sequencing and analysis of 454,787 uk biobank participants. Nature, 599(7886):628–634, 2021.

[26] Emre Guney, Jörg Menche, Marc Vidal, and Albert-László Barábasi. Network-based in silico drug efficacy screening. Nature communications, 7(1):1–13, 2016.

[27] Hai Fang, Hans De Wolf, Bogdan Knezevic, Katie L Burnham, Julie Osgood, Anna Sanniti, Alicia Lledó Lara, Silva Kasela, Stephane De Cesco, Jörg K Wegner, et al. A genetics-led approach defines the drug target landscape of 30 immune-related traits. Nature genetics, 51(7):1082–1091, 2019.

[28] Aidan MacNamara, Nikolina Nakic, Ali Amin Al Olama, Cong Guo, Karsten B Sieber, Mark R Hurle, and Alex Gutteridge. Network and pathway expansion of genetic disease associations identifies successful drug targets. Scientific reports, 10(1):1–11, 2020.

[29] Yingnan Han, Clarence Wang, Katherine Klinger, Deepak K Rajpal, and Cheng Zhu. An integrative network-based approach for drug target indication expansion. PloS one, 16(7):e0253614, 2021.

[30] Inigo Barrio-Hernandez, Jeremy Schwartzentruber, Anjali Shrivastava, Noemi del Toro, Qian Zhang, Glyn Bradley, Henning Hermjakob, Sandra Orchard, Ian Dunham, Carl A Anderson, et al. Network expansion of genetic associations defines a pleiotropy map of human cell biology. BioRxiv, 2021.

[31] Katja Luck, Dae-Kyum Kim, Luke Lambourne, Kerstin Spirohn, Bridget E Begg, Wenting Bian, Ruth Brignall, Tiziana Cafarelli, Francisco J Campos-Laborie, Benoit Charloteaux, et al. A reference map of the human binary protein interactome. Nature, 580(7803):402–408, 2020.

[32] Damian Szklarczyk, Annika L Gable, David Lyon, Alexander Junge, Stefan Wyder, Jaime Huerta-Cepas, Milan Simonovic, Nadezhda T Doncheva, John H Morris, Peer Bork, et al. String v11: protein–protein association networks with increased coverage, supporting functional discovery in genome-wide experimental datasets. Nucleic acids research, 47(D1):D607–D613, 2019.

[33] Patrick Deelen, Sipko van Dam, Johanna C Herkert, Juha M Karjalainen, Harm Brugge, Kristin M Abbott, Cleo C van Diemen, Paul A van der Zwaag, Erica H Gerkes, Evelien Zonneveld-Huijssoon, et al. Improving the diagnostic yield of exome-sequencing by predicting gene–phenotype associations using large-scale gene expression analysis. Nature communications, 10(1):1–13, 2019.

[34] Mikaela Koutrouli, Pau Piera Ĺındez, Robbin Bouwmeester, Lennart Martens, and Lars Juhl Jensen. Fava: High-quality functional association networks inferred from scrna-seq and proteomics data. bioRxiv, 2022.

[35] David S Wishart, Yannick D Feunang, An C Guo, Elvis J Lo, Ana Marcu, Jason R Grant, Tanvir Sajed, Daniel Johnson, Carin Li, Zinat Sayeeda, et al. Drugbank 5.0: a major update to the drugbank database for 2018. Nucleic acids research, 46(D1):D1074–D1082, 2018.

[36] Camilo Ruiz, Marinka Zitnik, and Jure Leskovec. Identification of disease treatment mechanisms through the multiscale interactome. Nature communications, 12(1):1–15, 2021.

[37] Anna Gaulton, Anne Hersey, Michał Nowotka, A Patricia Bento, Jon Chambers, David Mendez, Prudence Mutowo, Francis Atkinson, Louisa J Bellis, Elena Cibrián-Uhalte, et al. The chembl database in 2017. Nucleic acids research, 45(D1):D945–D954, 2017.

[38] Sharon L Freshour, Susanna Kiwala, Kelsy C Cotto, Adam C Coffman, Joshua F McMichael, Jonathan J Song, Malachi Griffith, Obi L Griffith, and Alex H Wagner. Integration of the drug– gene interaction database (dgidb 4.0) with open crowdsource efforts. Nucleic acids research, 49(D1):D1144–D1151, 2021.

[39] Damian Szklarczyk, Alberto Santos, Christian Von Mering, Lars Juhl Jensen, Peer Bork, and Michael Kuhn. Stitch 5: augmenting protein–chemical interaction networks with tissue and affinity data. Nucleic acids research, 44(D1):D380–D384, 2016.

[40] Luke J O’Connor, Armin P Schoech, Farhad Hormozdiari, Steven Gazal, Nick Patterson, and Alkes L Price. Extreme polygenicity of complex traits is explained by negative selection. The American Journal of Human Genetics, 105(3):456–476, 2019.

[41] Edward Mountjoy, Ellen M Schmidt, Miguel Carmona, Jeremy Schwartzentruber, Gareth Peat, Alfredo Miranda, Luca Fumis, James Hayhurst, Annalisa Buniello, Mohd Anisul Karim, et al. An open approach to systematically prioritize causal variants and genes at all published human gwas trait-associated loci. Nature Genetics, 53(11):1527–1533, 2021.

[42] Vincenzo Forgetta, Lai Jiang, Nicholas A Vulpescu, Megan S Hogan, Siyuan Chen, John A Morris, Stepan Grinek, Christian Benner, Dong-Keun Jang, Quy Hoang, et al. An effector index to predict target genes at gwas loci. Human Genetics, pages 1–17, 2022.

[43] Douglas W Yao, Luke J O’Connor, Alkes L Price, and Alexander Gusev. Quantifying genetic effects on disease mediated by assayed gene expression levels. Nature Genetics, 52(6):626–633, 2020.

[44] Ryan S Dhindsa, Oliver S Burren, Benjamin B Sun, Bram P Prins, Dorota Matelska, Eleanor Wheeler, Jonathan Mitchell, Erin Oerton, Ventzislava A Hristova, Katherine R Smith, et al. Influences of rare protein-coding genetic variants on the human plasma proteome in 50,829 uk biobank participants. bioRxiv, 2022.

[45] Alexander S Hauser, Sreenivas Chavali, Ikuo Masuho, Leonie J Jahn, Kirill A Martemyanov, David E Gloriam, and M Madan Babu. Pharmacogenomics of gpcr drug targets. Cell, 172(1-2):41–54, 2018.

[46] Evangelos Evangelou, Helen R Warren, David Mosen-Ansorena, Borbala Mifsud, Raha Pazoki, He Gao, Georgios Ntritsos, Niki Dimou, Claudia P Cabrera, Ibrahim Karaman, et al. Genetic analysis of over 1 million people identifies 535 new loci associated with blood pressure traits. Nature genetics, 50(10):1412–1425, 2018.

[47] Clare Bycroft, Colin Freeman, Desislava Petkova, Gavin Band, Lloyd T Elliott, Kevin Sharp, Allan Motyer, Damjan Vukcevic, Olivier Delaneau, Jared O’Connell, et al. The UK Biobank resource with deep phenotyping and genomic data. Nature, 562(7726):203–209, 2018.

[48] Matthias Wuttke, Yong Li, Man Li, Karsten B Sieber, Mary F Feitosa, Mathias Gorski, Adrienne Tin, Lihua Wang, Audrey Y Chu, Anselm Hoppmann, et al. A catalog of genetic loci associated with kidney function from analyses of a million individuals. Nature genetics, 51(6):957–972, 2019.

[49] Mitja I Kurki, Juha Karjalainen, Priit Palta, Timo P Sipilä, Kati Kristiansson, Kati Donner, Mary P Reeve, Hannele Laivuori, Mervi Aavikko, Mari A Kaunisto, et al. Finngen: Unique genetic insights from combining isolated population and national health register data. medRxiv, 2022.

[50] Cristen J Willer, Yun Li, and Gonçalo R Abecasis. Metal: fast and efficient meta-analysis of genomewide association scans. Bioinformatics, 26(17):2190–2191, 2010.

[51] Daniel Krefl and Sven Bergmann. Bergmannlab/pascalx: Pascalx v0.0.1 (v0.0.1), 2021. https://doi.org/10.5281/zenodo.4429922.

[52] UK10K, et al. The UK10K project identifies rare variants in health and disease. Nature, 526(7571):82, 2015.

[53] Zhihong Zhu, Futao Zhang, Han Hu, Andrew Bakshi, Matthew R Robinson, Joseph E Powell, Grant W Montgomery, Michael E Goddard, Naomi R Wray, Peter M Visscher, et al. Integration of summary data from gwas and eqtl studies predicts complex trait gene targets. Nature genetics, 48(5):481–487, 2016.

[54] Gibran Hemani, Kate Tilling, and George Davey Smith. Orienting the causal relationship between imprecisely measured traits using GWAS summary data. PLoS genetics, 13(11):e1007081, 2017.

[55] THE GTEX CONSORTIUM. The GTEx Consortium atlas of genetic regulatory effects across human tissues. Science, 369(6509):1318–1330, 2020.

[56] Jon Chambers, Mark Davies, Anna Gaulton, Anne Hersey, Sameer Velankar, Robert Petryszak, Janna Hastings, Louisa Bellis, Shaun McGlinchey, and John P Overington. Unichem: a unified chemical structure cross-referencing and identifier tracking system. Journal of cheminformatics, 5(1):1–9, 2013.

[57] Steffen Durinck, Paul T. Spellman, Ewan Birney, and Wolfgang Huber. Mapping identifiers for the integration of genomic datasets with the r/bioconductor package biomart. Nature Protocols, 4:1184–1191, 2009.

[58] Andrew Nightingale, Ricardo Antunes, Emanuele Alpi, Borisas Bursteinas, Leonardo Gonzales, Wudong Liu, Jie Luo, Guoying Qi, Edd Turner, and Maria Martin. The proteins api: accessing key integrated protein and genome information. Nucleic acids research, 45(W1):W539–W544, 2017.

[59] Xavier Robin, Natacha Turck, Alexandre Hainard, Natalia Tiberti, Frédérique Lisacek, Jean-Charles Sanchez, and Markus Mü ller. proc: an open-source package for r and s+ to analyze and compare roc curves. BMC bioinformatics, 12(1):1–8, 2011.

[60] Hanghang Tong, Christos Faloutsos, and Jia-Yu Pan. Fast random walk with restart and its applications. In Sixth international conference on data mining (ICDM’06), pages 613–622. IEEE, 2006.

[61] Gabor Csardi, Tamas Nepusz, et al. The igraph software package for complex network research. InterJournal, complex systems, 1695(5):1–9, 2006.

