## Supplementary Figures for "Multi-layered genetic approaches to identify approved drug targets"

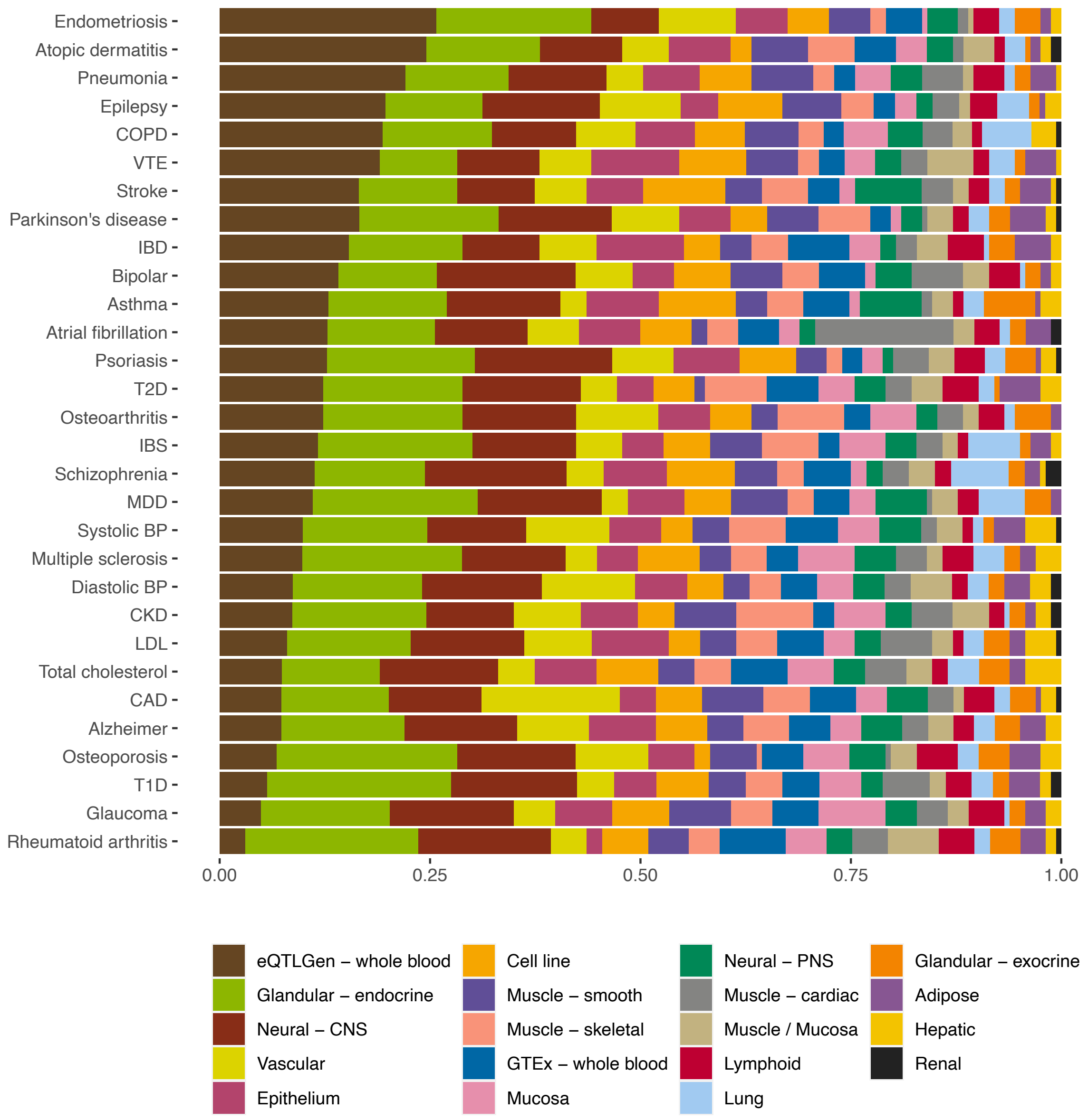


**Supplementary Figure 1**. Proportion of genes mapped to a particular tissue category in the tissue-wide expression quantitative trait locus (eQTL)-genome-wide association analysis (GWAS) analysis. For each gene, the tissue with the lowest Mendelian randomization (MR) p-value was selected. Tissue category belonging are shown in Table S4 and numerical proportion values in Table S5.

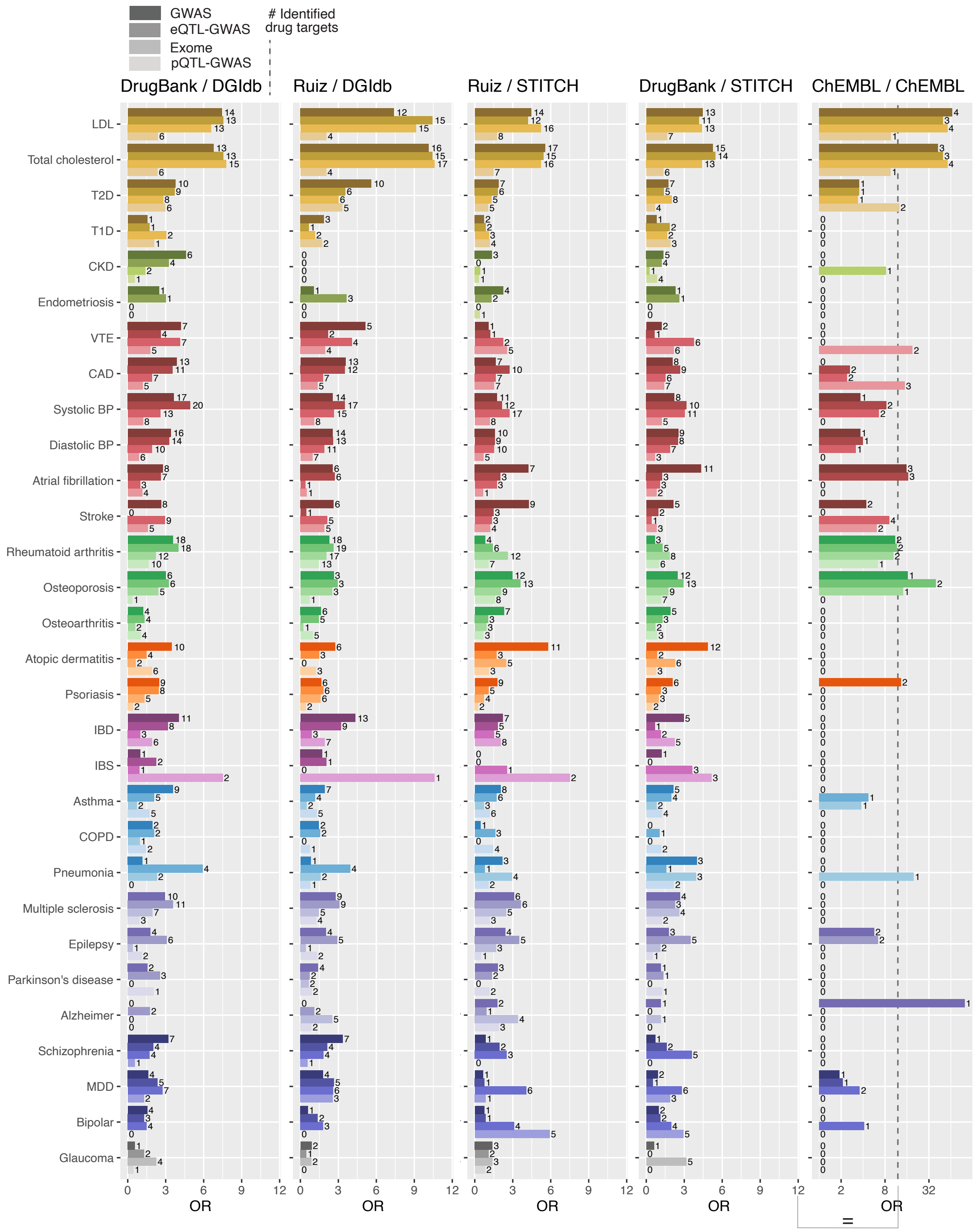


**Supplementary Figure 2**. Enrichment for drug target genes across drug databases. Barplots with odds ratios (ORs) calculated from Fisher's exact tests between drug target genes and prioritized genes for the four tested methods and thirty traits. Prioritized genes were defined as the top 1% percentile of the GWAS, eQTL-GWAS and Exome methods, and 5% of the pQTL-GWAS method. Drug target genes were defined by the drug database combinations (drug-indication and drug-target links) shown in the title of each barplot. Only drug target genes that could be tested by the respective method were considered. The number on the right of each bar indicates the number of identified drug target genes. In the barplot corresponding to the ChEMBL/ChEMBL database, the x-axis is log-transformed and therefore ORs of 0 (i.e., no identified drug target) were set to 1.

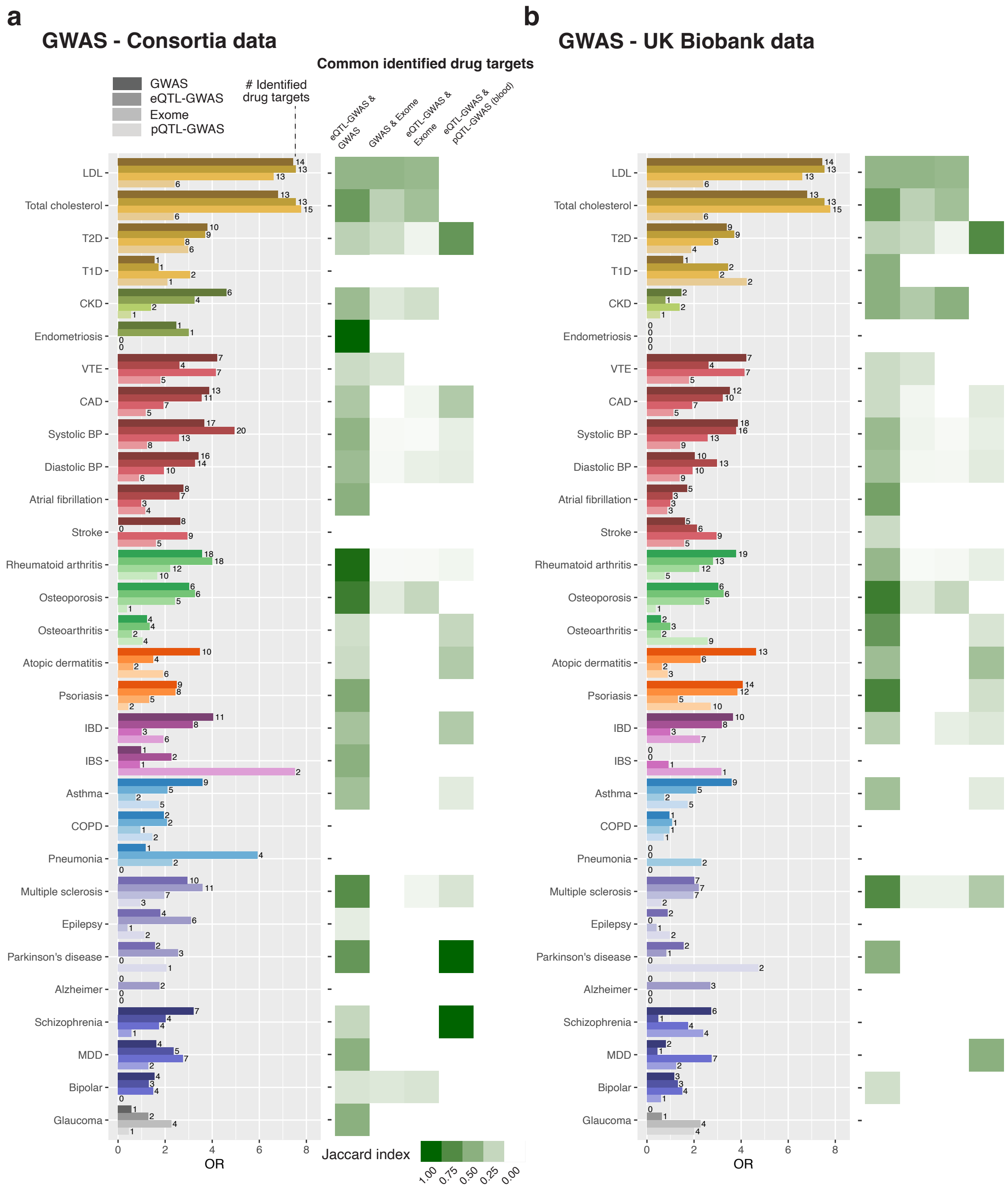


**Supplementary Figure 3**. Comparing the utilization of consortia GWAS and UK Biobank GWAS data in the enrichment analysis for drug target genes. **a** Enrichment analysis using consortia GWAS summary statistics in the GWAS, eQTL-GWAS and pQTL-GWAS methods. **b** Enrichment analysis using UKBB GWAS summary statistics in the GWAS, eQTL-GWAS and pQTL-GWAS methods. The Exome analysis is only performed on UK Biobank data. Left: Barplot with odds ratios (ORs) calculated from Fisher's exact tests between drug target genes and prioritized genes for the four tested methods and thirty traits. Prioritized genes were defined as the top 1% percentile of the GWAS, eQTL-GWAS and Exome methods, and 5% of the pQTL-GWAS method. Drug target genes were defined from the DrugBank and DGIdb databases, and only drug target genes that could be tested by the respective method were considered. The number on the right of each bar indicates the number of identified drug target genes. Right: Overlap of identified drug target genes between pairs of methods quantified through the Jaccard index. The blood-only eQTL-GWAS gene prioritization method was used for the comparison with the pQTL-GWAS method.

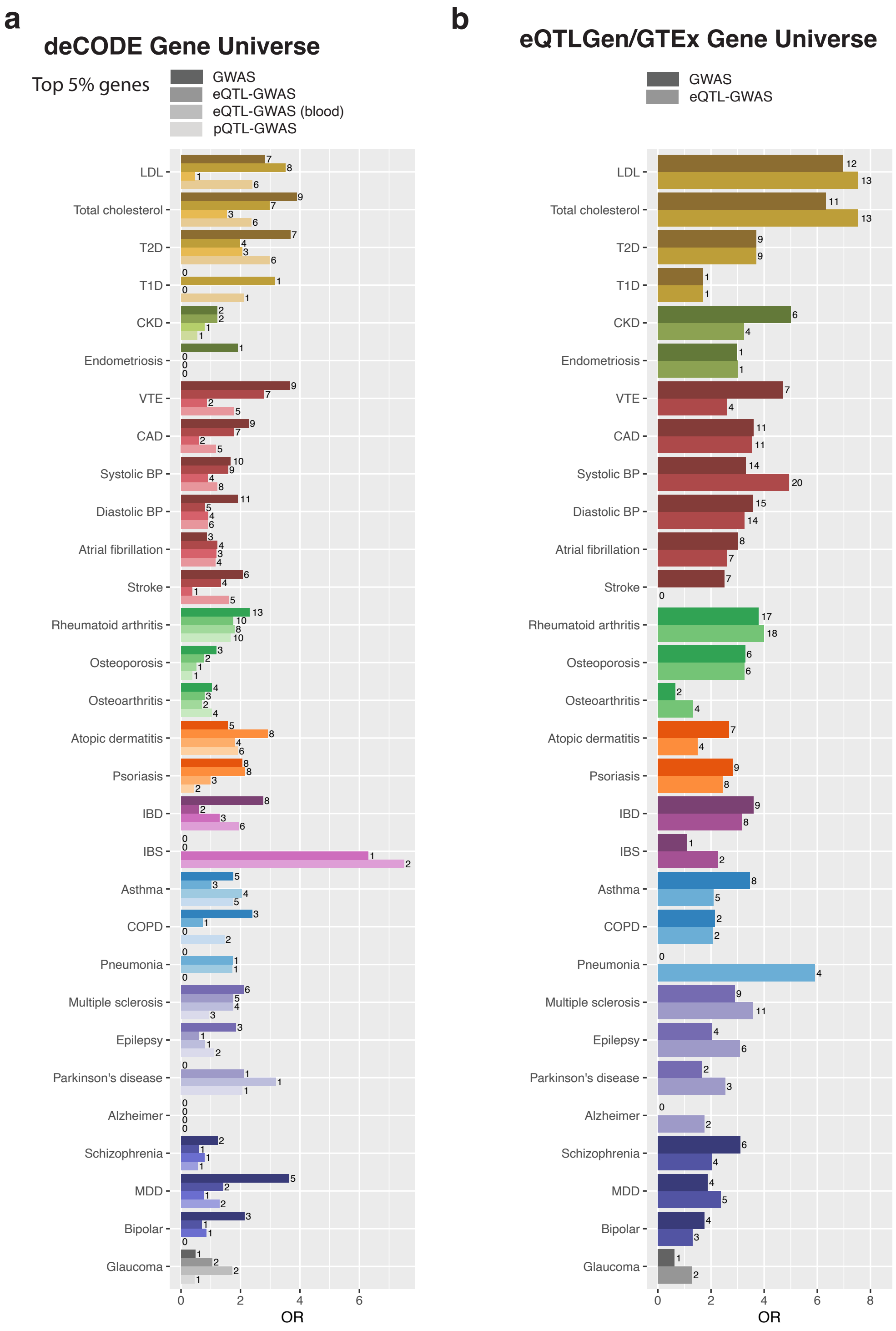


**Supplementary Figure 4**. Enrichment for drug target genes using the same background genes. **A** Enrichment analysis was performed by subsetting the gene universe of the GWAS and eQTL-GWAS (tissue-wide and whole blood only) methods to the genes available in the deCODE study (i.e., proteins used in the pQTL-GWAS analysis). **b** Enrichment analysis was performed by subsetting the gene universe of the GWAS method to the genes available in the tissue-wide eQTL-GWAS analysis. Both plots show barplots with odds ratios (ORs) calculated from Fisher's exact tests between drug target genes and prioritized genes for the four tested methods and thirty traits. Drug target genes were defined from the DrugBank and DGIdb databases, and only drug target genes that could be tested by the respective method were considered. The number on the right of each bar indicates the number of identified drug target genes.

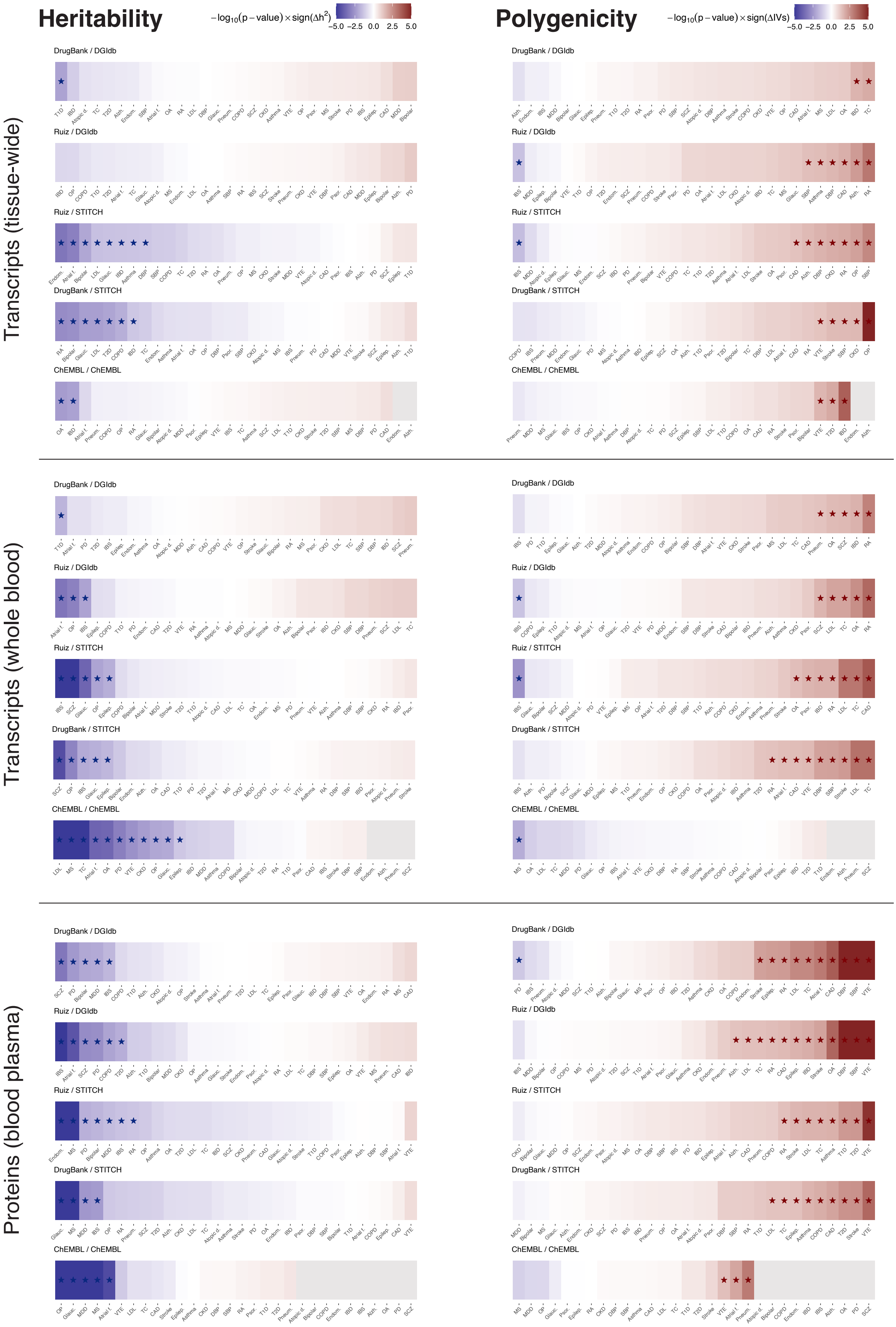
Heritability
$-\log_{10}(p-\text{value}) \times \text{sign}(\Delta h^2)$
−5.0 −2.5 0.0 2.5 5.0
Polygenicity
$-\log_{10}(p-\text{value}) \times \text{sign}(\Delta \text{IVs})$
−5.0 −2.5 0.0 2.5 5.0
Transcripts (tissue-wide)
Transcripts (whole blood)
Proteins (blood plasma)
DrugBank / DGIdb
Ruiz / DGIdb
Ruiz / STITCH
DrugBank / STITCH
ChEMBL / ChEMBL

**Supplementary Figure 5**. Difference in heritability and polygenicity of drug target compared to non-drug target measured transcript and protein levels. For each trait, the difference in heritability was calculated through a two-sided t-test. The difference in polygenicity was calculated through a two-sided Wilcoxon test. When the difference was negative (i.e., drug target genes were less heritable or less polygenic), the $-\log_{10}$(p-value) is plotted in blue, otherwise in red. Traits for which the difference was nominally significant (p-value < 0.05), are indicated with a star. If less than three drug target genes could be tested for a trait, a grey box is plotted.

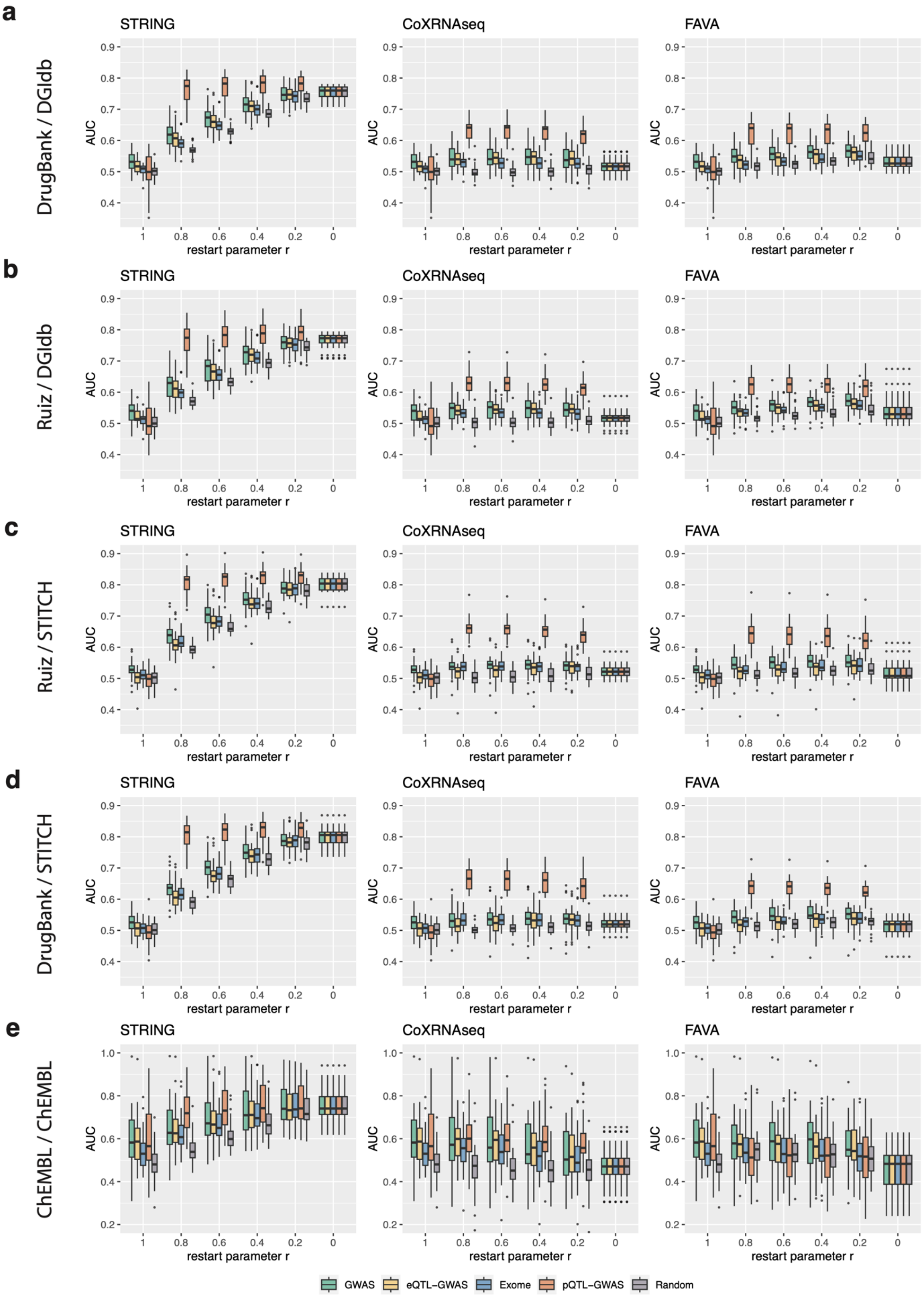


**Supplementary Figure 6.** Effect of network diffusion to prioritize drug target genes across drug databases (AUC values). Boxplots showing the area under the receiver operating characteristic curve (AUC) values for each network type (STRING, CoXRNAseq and FAVA) and method at different restart parameter values *r*. AUC values were calculated for each of the thirty traits, and drug target genes were defined by the respective drug database combination (drug-indication and drug-target links, **a**-**e**). The boxplots bound the 25th, 50th (median, centre), and the 75th quantile. Whiskers range from minima (Q1 – 1.5 • IQR) to maxima (Q3 + 1.5 • IQR) with points above or below representing potential outliers.

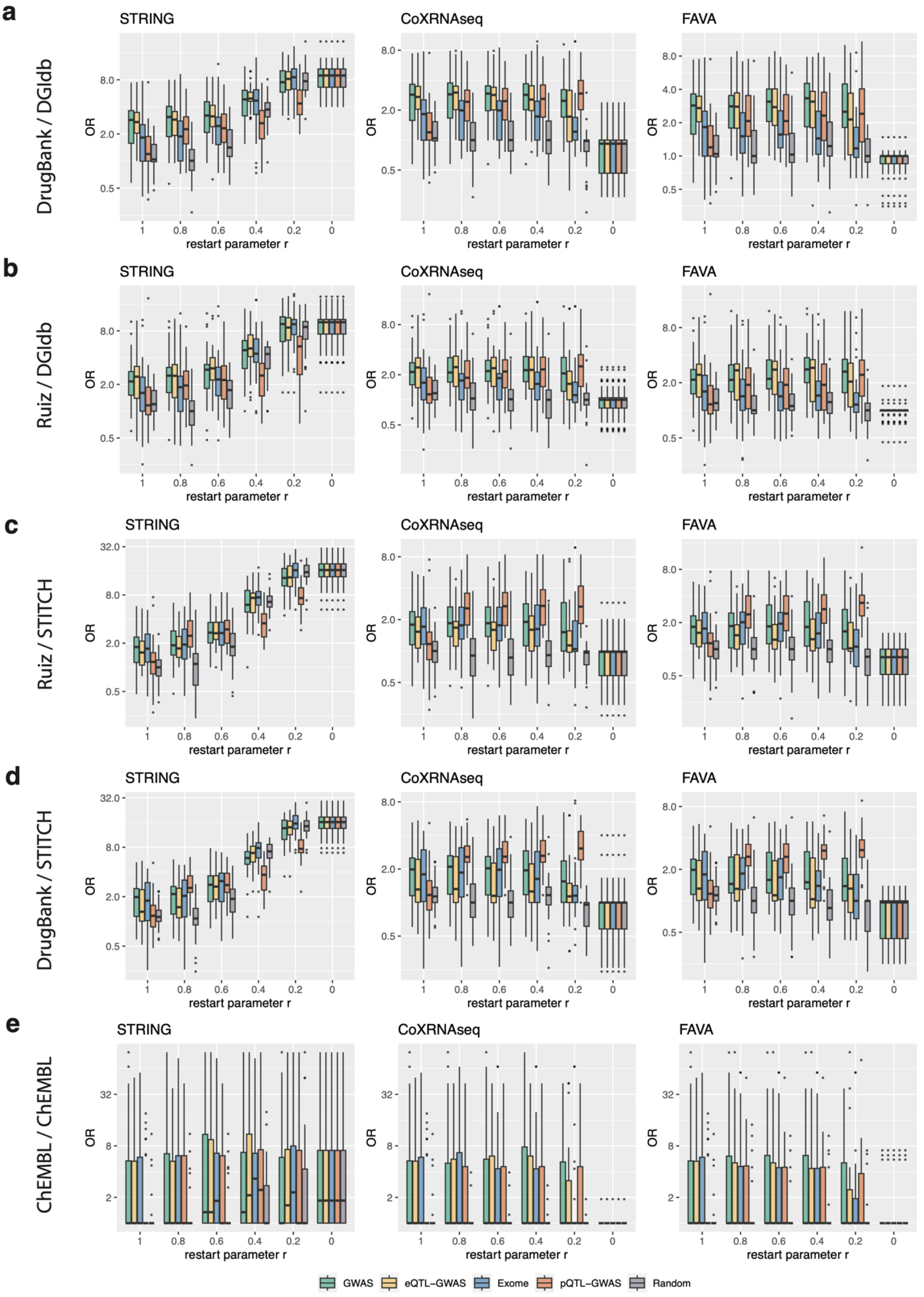


**Supplementary Figure 7**. Effect of network diffusion to prioritize drug target genes across drug databases (ORs). Odds ratios (ORs) between prioritized genes (top 1%) and drug target genes for each network type (STRING, CoXRNAseq and FAVA) and method at different restart parameter values *r*.

Drug target genes were defined by the respective drug database combination (drug-indication and drug-target links, **a**-**e**). The OR was set to 1 for traits with no identified drug target genes. The boxplots bound the 25th, 50th (median, centre), and the 75th quantile. Whiskers range from minima (Q1 – 1.5 • IQR) to maxima (Q3 + 1.5 • IQR) with points above or below representing potential outliers.

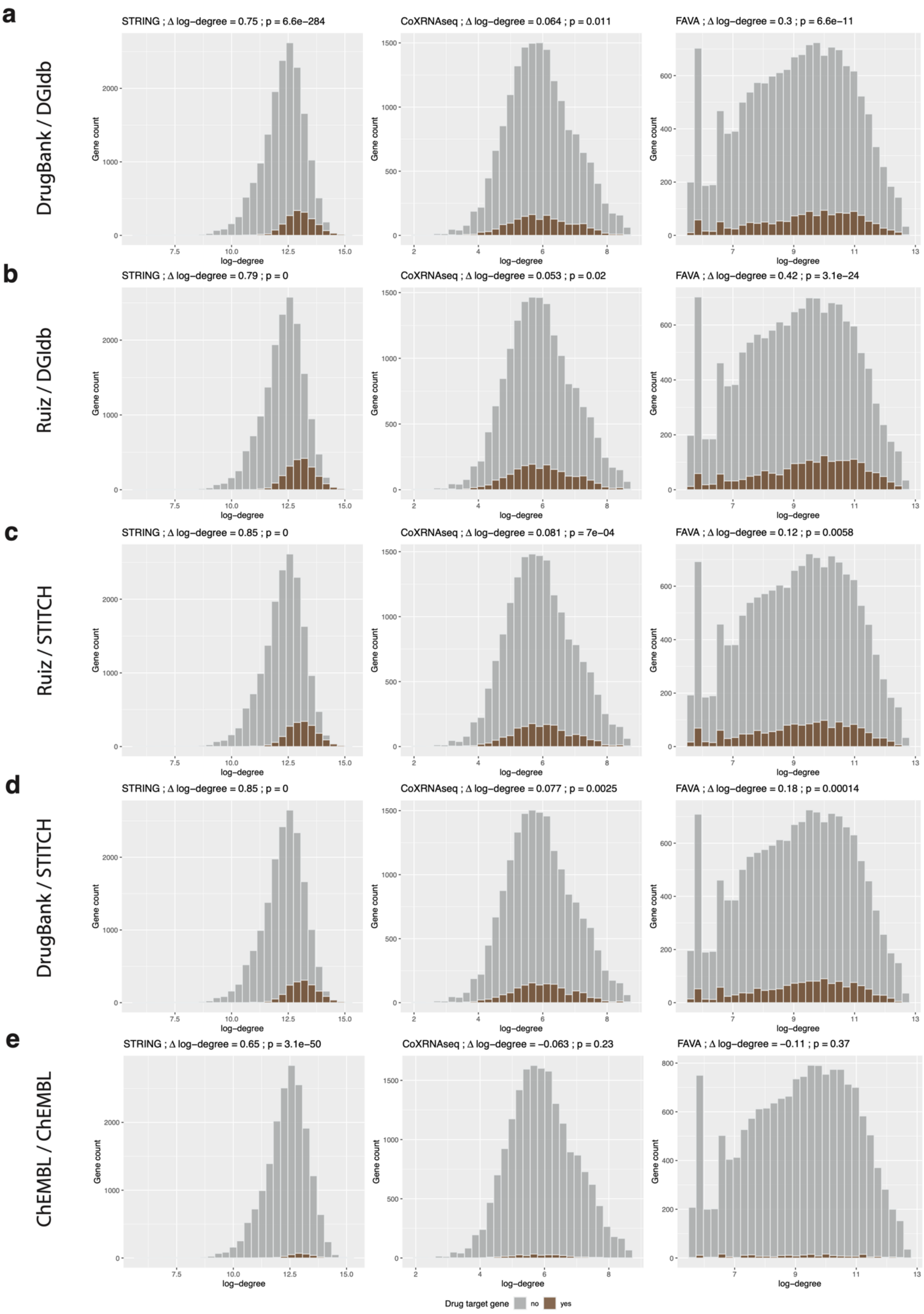
a
DrugBank / DGIdb
STRING ; Δ log−degree = 0.75 ; p = 6.6e−284
CoXRNAseq ; Δ log−degree = 0.064 ; p = 0.011
FAVA ; Δ log−degree = 0.3 ; p = 6.6e−11
b
Ruiz / DGIdb
STRING ; Δ log−degree = 0.79 ; p = 0
CoXRNAseq ; Δ log−degree = 0.053 ; p = 0.02
FAVA ; Δ log−degree = 0.42 ; p = 3.1e−24
c
Ruiz / STITCH
STRING ; Δ log−degree = 0.85 ; p = 0
CoXRNAseq ; Δ log−degree = 0.081 ; p = 7e−04
FAVA ; Δ log−degree = 0.12 ; p = 0.0058
d
DrugBank / STITCH
STRING ; Δ log−degree = 0.85 ; p = 0
CoXRNAseq ; Δ log−degree = 0.077 ; p = 0.0025
FAVA ; Δ log−degree = 0.18 ; p = 0.00014
e
ChEMBL / ChEMBL
STRING ; Δ log−degree = 0.65 ; p = 3.1e−50
CoXRNAseq ; Δ log−degree = −0.063 ; p = 0.23
FAVA ; Δ log−degree = −0.11 ; p = 0.37
Gene count
log−degree
Drug target gene
no
yes

**Supplementary Figure 8**. Histograms showing the degree distribution of drug target genes and non-drug target genes in each network across drug databases (drug-indication and drug-target links, **a**-**e**). The difference in log-degree and the p-values from two-sided t-tests are shown in the title.
